# Hyperreflective foci, OCT progression indicators in age-related macular degeneration, include transdifferentiated retinal pigment epithelium

**DOI:** 10.1101/2021.04.26.21256056

**Authors:** Dongfeng Cao, Belinda Leong, Jeffrey D. Messinger, Deepayan Kar, Thomas Ach, Lawrence A. Yannuzzi, K. Bailey Freund, Christine A. Curcio

**Affiliations:** Department of Ophthalmology and Visual Sciences, University of Alabama at Birmingham School of Medicine; Birmingham, AL, USA; Vitreous Retina Macula Consultants of New York; New York, NY, USA; LuEsther T. Mertz Retinal Research Center, Manhattan Eye, Ear and Throat Hospital; New York, NY, USA; Retina Associates; Sydney, NSW, Australia; Department of Ophthalmology, University Hospital Bonn; Bonn, Germany; Department of Ophthalmology, New York University, Grossman School of Medicine; New York, NY, USA

**Author notes:** These authors contributed equally to this work (Dongfeng Cao, Belinda Leong). Previous meeting presentations: annual meeting of the Association for Research in Vision and Ophthalmology, 2019 (clinical imaging) and 2020 (laboratory study).

**Keywords:** age-related macular degeneration, hyperreflective foci, retinal pigment epithelium, optical coherence tomography, immunohistochemistry, retinoids, immune markers, epithelial-mesenchyme transition

## Abstract

**Purpose:** By optical coherence tomography (OCT) imaging, hyperreflective foci (HRF) indicate progression risk for advanced age-related macular degeneration (AMD) and are in part attributable to ectopic retinal pigment epithelium (RPE). We hypothesized that ectopic RPE are molecularly distinct from in-layer cells and that their cross-retinal course follows Müller glia.

**Method:** In clinical OCT (61 eyes, 44 AMD patients, 79.4 ± 7.7 years; 29 female; follow-up = 4.7 ± 0.9 years) one HRF type, RPE plume, was reviewed. Twenty eyes of 20 donors characterized by *ex vivo* OCT were analyzed by histology (normal, 4; early/intermediate AMD, 7; geographic atrophy (GA), 6; 3 neovascular AMD (nvAMD). Cryosections were stained with antibodies to retinoid (RPE65, CRALPB) and immune (CD68, CD163) markers. In published RPE cellular phenotypes, red immunoreactivity was assessed semi-quantitatively by one observer (none, some cells, all cells).

**Results:** Trajectories of RPE plume and cellular debris paralleled Müller glia, whether ordered or subsident, near atrophy borders. RPE corresponding to HRF lost immunoreactivity for retinoid markers and gained immunoreactivity for immune markers. Aberrant immunoreactivity appeared in individual in-layer RPE cells and extended to all abnormal phenotypes. Müller glia remained CRALBP-positive. Plume cells approached and contacted retinal capillaries.

**Conclusion:** Gain- and loss-of-function starts with individual in-layer RPE cells and extends to all abnormal phenotypes. Down-regulated RPE retinoid handling may impair rod vision while Müller glia sustain cone vision. Evidence for RPE transdifferentiation, possibly due to ischemia, supports a proposed process of epithelial-mesenchyme transition. Data can propel new biomarkers and therapeutic strategies for AMD.

**Précis:** Hyperreflective foci (HRF) are OCT progression risk indicators in age-related macular degeneration. Abnormal RPE cells including some that correspond to HRF lose immunoreactivity for retinoid markers and gain immunoreactivity for immune markers, indicating molecular transdifferentiation.

## INTRODUCTION

Age-related macular degeneration (AMD) causes vision loss in older persons worldwide. ^1^ Neovascular complications are managed medically, ^2^ whereas the underlying and prevalent atrophic (“dry”) disease is not yet treatable. AMD affects vertically integrated cellular layers formed by light-sensing photoreceptors and support cells (retinal pigment epithelium (RPE), choriocapillaris, Müller glia). Characteristic extracellular deposits (drusen, subretinal drusenoid deposits) form between photoreceptors and their choroidal and retinal blood supplies. ^3^

AMD is increasingly understood via clinical optical coherence tomography (OCT). This technology provides depth-resolved and registered images of retinal and choroidal tissue layers ^4^ that resemble histology and can be followed over time. ^5^ Not only are RPE cells central to, and impacted by, both atrophic and neovascular AMD, they can be uniquely monitored in vivo by OCT. RPE health is also an OCT readout of the less visible Bruch’s membrane and choriocapillaris. Each RPE cell has >1400 organelles ^6^ capable of generating OCT reflectivity. Further, as neovascular AMD is managed clinically, many OCT images of at-risk eyes are captured. A progression sequence for drusen-initiated atrophy supported by OCT and histology is age-related deterioration of choriocapillaris and Bruch’s membrane preventing egress of wastes to the circulation, growth of drusen with death or migration of overlying RPE, and reactive gliosis. ^5^

Longitudinal studies and post-hoc analysis of clinical trial imaging datasets revealed OCT biomarkers for progression from intermediate AMD to geographic atrophy. Tissue damage in atrophy is severe and possibly irreversible. ^7^ Currently the only imaging outcome accepted by the U.S. Food and Drug Administration for AMD trials is slowing the expansion of atrophy. Identifying outcome measures earlier in disease progression is the goal of an international consensus group. ^5, 8-10^ Of OCT biomarkers, hyperreflective foci (HRF) within the neurosensory retina confer the largest progression risk. ^11-14^ Our histologic survey of RPE phenotypes in AMD, ^15-17^ confirmed in clinically characterized eyes, ^15, 18^ identified two RPE phenotypes corresponding to HRF. These are “sloughed” cells in the subretinal space and “intraretinal” cells within the neurosensory retina. In a hypothetical progression sequence (Fig. 1), morphologically distinct cells deploy along separate pathways of cell fate. These include death, migration, and transdifferentiation to other cell types, without intervening dedifferentiation and proliferation. ^19^ Due to migration and death-in-place of RPE, the layer disintegrates, scattering fully pigmented and nucleated cells (“dissociated”) across atrophic areas.

**Figure 1.**
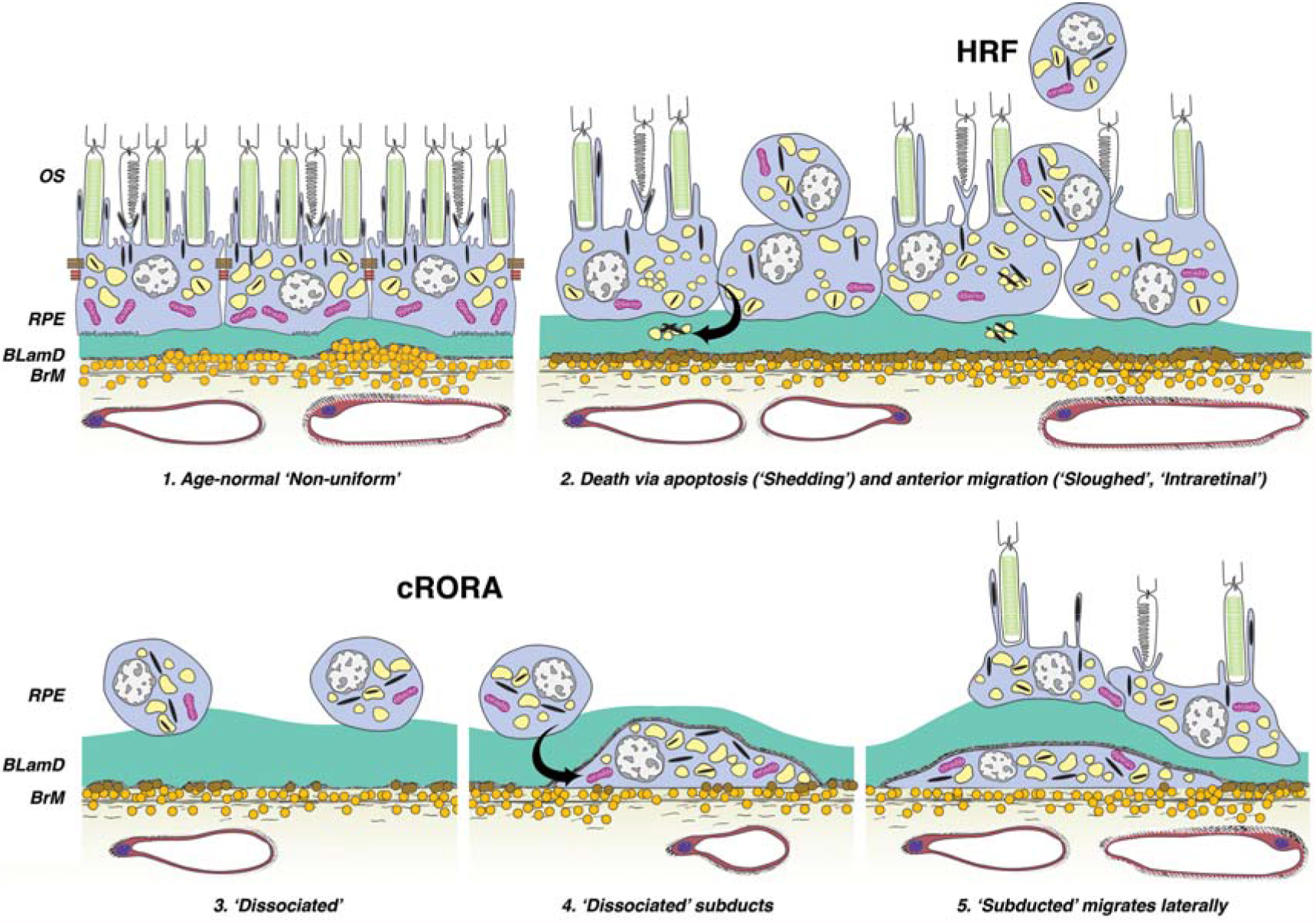
Hypothesis for complete retinal pigment epithelium (RPE) and outer retinal atrophy (cRORA) in AMD^9^ based on histology and OCT. This schematic for non-neovascular AMD is adapted from Zanzottera et al. ^44^ Six morphologic phenotypes of RPE from a system of 15 ^27^ are shown. (**1**) Age-normal “nonuniform” RPE overlies Bruch’s membrane (BrM), which has abundant lipoprotein particles (yellow). Basal laminar deposit (BLamD, green) is thickened basement membrane material between the RPE cell body and native RPE basal lamina. RPE organelles are melanosomes (black), lipofuscin (yellow), and mitochondria (pink). ^6^ (**2**) Anteriorly migrated RPE (“sloughed” and “intraretinal”) manifest clinically as HRF. “Shedding” cells release organelle clusters ^15^ (arrow) basally into BLamD, thought to represent apoptosis. (**3**) Due to cell death and migration, the RPE layer disintegrates. “Dissociated” RPE are fully pigmented, nucleated cells scattered in the atrophic zone. BLamD persists after RPE death. (**4)** “Subducted” cells containing RPE organelles, appearing to originate from Dissociated cells (arrow), are flattened against BrM. (**5**) “Subducted” cells migrate outside the atrophic area.

Some cells appear to dive below the intact layer and migrate outside the atrophic area (“subducted”). ^16^ In neovascular AMD, lipid filled non-RPE cells also correlated to HRF ^18, 20^ and may represent microglia, macrophages, or both.

We postulated that one HRF form called “RPE plume” represents cells crossing the retina within the Henle fiber layer (HFL), which varies in thickness and angle with retinal position (Supplementary Fig. 1). ^17^ We herein provide a clinical OCT description for this feature in patients with AMD and show RPE-derived debris in relation to the cross-retinal course of Müller glia. To probe the identity of HRF and test whether they include transdifferentiated RPE, we used immunohistochemistry of human donor eyes to show loss of retinoid processing and gain of immunologic function in all abnormal RPE phenotypes. Widespread RPE transdifferentiation, consistent with a proposed process of epithelial-mesenchyme transition, ^21^ supports a strategy of visualizing and targeting pathology earlier in AMD progression than atrophy.

## MATERIALS AND METHODS

### Compliance

Study of a clinical cohort and use of human tissue samples was approved by institutional review at Vitreous Macula Retina and University of Alabama at Birmingham (IRB-150910011), respectively, and complied with the Health Insurance Portability and Accountability Act of 1996 and the Declaration of Helsinki.

### Patient selection, clinical image capture, and image analysis

Consensus nomenclature was used for reflective bands of OCT. ^22^ RPE+BL band refers to the RPE and its basal lamina atop drusen or other material in the sub-RPE-BL space. This space is between RPE-BL and inner collagenous layer of Bruch’s membrane. ^23^

To analyze RPE plume natural history, patients with non-neovascular AMD on OCT imaging seen by specialists (authors KBF, LAY) at a tertiary private retinal practice were retrospectively reviewed (author BL). Inclusion criteria included age >50 years, >12 months of follow-up with tracked OCT B-scans at an inter-scan spacing of ≤250 μm, and maximum follow-up interval of 6 months, and at least one eye containing HRF. HRF were defined as features with reflectivity like that of the RPE, separated from the RPE layer (i.e., discrete HRF), and thought to originate from the RPE layer. Reduced RPE+BL thickness in a scan subsequent to HRF appearance was considered evidence for RPE origin. Patients with diabetic retinopathy, retinal vascular disease, previous vitreoretinal surgery, laser treatment or ocular inflammation were excluded. Eyes were evaluated at baseline by presence of AMD lesions including soft and cuticular drusen, subretinal drusenoid deposits, and acquired vitelliform lesions ^24^). If color fundus photographs were available, pigmentary changes were also considered. Presence of drusenoid pigment epithelial detachment ^25^ and complete RPE and outer retinal atrophy (cRORA) ^9^ was noted.

All patients underwent spectral domain OCT and near-infrared reflectance imaging (Spectralis HRA + OCT, Heidelberg Engineering, Heidelberg, Germany) with eye tracking enabled. The OCT protocol comprised 20° horizontal raster line scans centered on the fovea (19 to 49 B-scans per eye; scan spacing at 115 to 250 µm, automatic real-time averaging between 4 and 23).

Consecutive tracked OCT and near-infrared reflectance images were analyzed over time and with reference to the foveal center, from the date of HRF appearance. Plume status over time was recorded as no change, change to a different morphology, formation of a discrete HRF as defined above, and dissolution. Plume direction towards the OPL was noted as congruent or incongruent with normal HFL architecture (Supplementary Fig. 1), i.e., parallel or non-parallel with fibers. To evaluate plumes as progression indicators, expansion of cRORA and drusen growth and collapse were also noted.

### Overview of laboratory studies

To elucidate the molecular repertoire of RPE phenotypes corresponding to HRF, we obtained human donor eyes with and without AMD and performed immunohistochemistry. We used antibodies for retinoid- and immune-related proteins ^26^ (Supplementary Table 1) and positive and negative controls in human tissues (Supplementary Fig. 2). Cryosections were processed with an avidin-biotin-peroxidase immunohistochemistry detection system and a red reaction product. We checked the immunolabeling of all RPE phenotypes ^27^ (Supplementary Table 2) using bright field microscopy for direct observation of cellular pigmentation and status of surrounding tissue.

### Donor eye acquisition, *ex vivo* imaging, AMD diagnosis

Whole eyes were obtained from deceased human donors (≥ 80 years of age, white, non-diabetic, and ≤6 hours death-to-preservation) to the Advancing Sight Network (Birmingham AL USA) during 2016-17. Eye health history was not available. After anterior segment removal, eyes were preserved in 4% buffered paraformaldehyde, screened for AMD using *ex vivo* multimodal imaging including OCT ^17^ (details in Results). Tissues were stored in 1% buffered paraformaldehyde at 4°C until used.

Rectangles of full-thickness eye wall (5×8 mm) including fovea and optic nerve were dissected. To prevent ice crystals, tissue was cryoprotected in ascending concentrations of sucrose in phosphate buffered saline (PBS) then ascending concentrations of tissue embedding medium and sucrose buffer (HistoPrep, Thermo Fisher Scientific, #SH75-1250D, Rockford IL USA); 4 parts sucrose buffer and 1 part medium then 2:1 sucrose: medium. Tissue was placed into a cryomold (Electron Microscopy Sciences, #70176-10, Fort Washington PA USA) with embedding medium, oriented, frozen in liquid nitrogen, and stored at −80°C. Tissues were sectioned at 12 µm thickness on a cryostat (Leica CM3050 Vashaw Scientific, Norcross GA USA), air dried at 37°C overnight, and stored at −80°C until used.

For AMD diagnosis and imaging-histology correlation, cryosections through the fovea and perifovea (recognized by HFL) were stained with periodic acid Schiff hematoxylin to highlight Bruch’s membrane and sub-RPE deposits. One stained section per slide was scanned with a 20X objective and a robotic microscope stage (Olympus VSI 120, CellSens; Olympus, Center Valley PA), scaled to tissue units, and centered on the fovea or vertical meridian (where Henle fibers diverge) using a custom plugin for FIJI (https://imagej.nih.gov/ij/download.html). *Ex vivo* OCT B-scans and scanned whole sections were matched for major landmarks (e.g., overall tissue contour, foveal center, large vessels, individual pathologies). For detailed examination, some sections were scanned with a 60X oil immersion objective (numerical aperture = 1.4). For figures, images were adjusted to maximize the intensity histogram for contrast and white balance and made into composites (Photoshop CS6, Adobe Systems, USA).

### Immunohistochemistry and evaluation

Detailed methods for immunohistochemistry are available in the Supplementary Materials.

Glass slides were scanned using the system described above and 20x and 40x objectives. RPE phenotype was categorized using the Project MACULA grading system. ^15, 16^ (Supplementary Table 2). In each stained section (1 per antibody per eye), immunoreactivities for each RPE phenotype were scored semi-quantitatively by a single observer (author DC after training by author CAC) as 0-not stained, 1-some stained and 2-all stained. These scores were converted to percentages using by total scores of eyes for each phenotype divided by maximum score (2 x number of eyes with the phenotype) times 100. Each phenotype was seen in at least 3 eyes.

## RESULTS

### Clinical imaging

Four morphologies of RPE plume were defined (Fig 2). Curvilinear plumes (Fig. 2A) appear as hyperreflective features of constant width, above the RPE layer with or without drusen, and pointing towards the outer plexiform layer. Relative to curvilinear, comma plumes (Fig. 2B) are similarly reflective, with a wider base at the RPE layer and tapering anteriorly. Like curvilinear, thread plumes (Fig. 2C), previously seen in inherited retinopathies, ^28^ are of uniform width but are thinner. A vitelliform plume (Fig. 2D) is thick at its base, typically an acquired vitelliform lesion (autofluorescent and yellow subretinal extracellular material), ^24^ with an anterior taper that widens over time.

**Figure 2.**
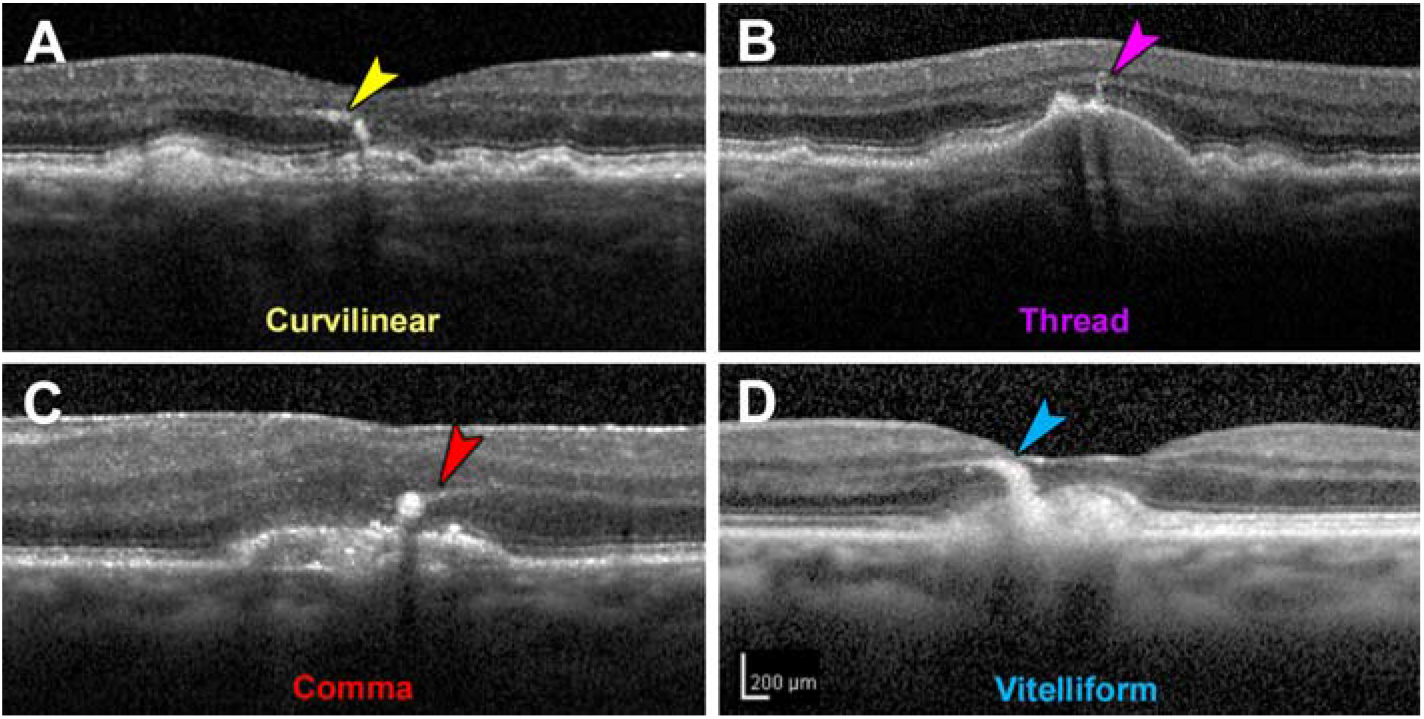
Morphology of retinal pigment epithelium plumes. Examples from two eyes of one patient. (**A**) “Curvilinear” consists of hyperreflective structures of consistent width, often arising from a soft druse and aiming towards the outer plexiform layer. (**B**) “Thread” has a uniform and thin width, compared to Curvilinear. (**C**) “Comma” has a wide base adjacent to the RPE layer and an anterior taper. (**D**) “Vitelliform” is thick and arises from an acquired vitelliform lesion. ^24^

To observe plume lifecycles and intraretinal trajectories, 44 patients with non-neovascular AMD (61 eyes, 29 females, 79.4 years (standard deviation 7.7 years)) with at least one HRF were enrolled (Supplementary Table 3). In 29 eyes of 21 patients (15 female), 129 plumes were localized and followed for characteristics including presence of cRORA and iRORA). ^5, 9^ Mean follow-up duration was 4.67 years (standard deviation 0.09 years). As shown in Supplementary Fig. 3A-B, plumes obliquely crossing horizontally oriented OCT B-scans appeared as small reflective dots that were connected by inspecting consecutive scans. Over time individual plumes evolved considerably (Fig. 3), converting from one morphology to another (Fig. 3B,D,E) or to discrete HRF as defined above (Fig. 3C). They also disappeared with (Fig. 3A,B,D) or persisted after (Fig. 3A, B, C, D) druse collapse to atrophy. One plume preceded druse appearance (Fig. 3E).

**Figure 3.**
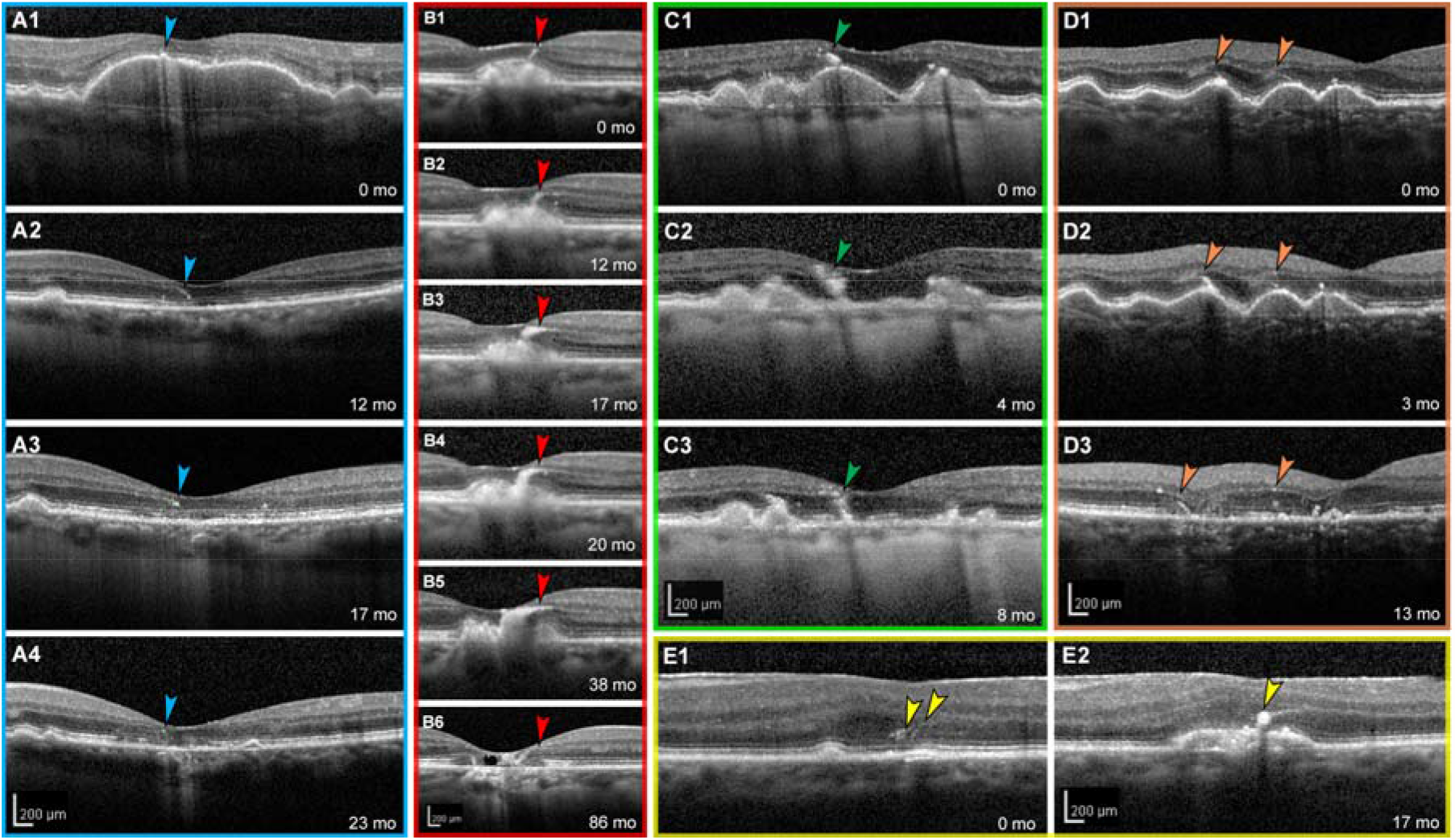
Retinal pigment epithelium plumes evolve over time. (**A** to **E**) Tracked optical coherence tomography B-scans of 5 different eyes with AMD are shown with individualized colored frames and scale bars. Numbers in the lower right corners indicate time elapsed in months since the baseline visit at 0 months. (**A1** to **A4**) Hyperreflective foci (HRF) overlie a drusenoid pigment epithelial detachment that collapsed at 12 months. HRF evolve into a comma plume before disappearing into complete outer retinal atrophy (cORA ^9^). (**B1** to **B6**) A curvilinear plume evolves into a vitelliform plume, which collapses, leaving atrophy and degenerative cysts. (**C1** to **C3)** A comma-type plume evolves into HRF upon drusen collapse. (**D1** to **D3**) Of two thread plumes at baseline, one (left) becomes curvilinear at 3 months and persists through drusen collapse and *incomplete* RPE and outer retinal atrophy at 13 months. Thread plume at the right has disappeared at 13 months. (**E1** to **E2**) Two thread plumes evolve into a comma plume as an RPE elevation appears. This eye also has an epiretinal membrane.

Curvilinear and thread plumes were the most common morphologies at baseline and the longest lasting over follow up (Supplementary Fig. 4A). Plume number and dynamism (i.e., appearance, disappearance, and conversions) could be striking (Supplementary Fig. 4B). Of 129 plumes at baseline (Supplementary Table 4), 73 (56.6%) converted to another plume morphology or discrete HRF during follow-up, with 38 disappearing and 35 remaining; 56 (43.4%) plumes retained the original morphology.

Most plumes (101/129, 78.3%) were geometrically congruent with the local trajectory of the normal HFL (Supplementary Table 5), especially temporal to the fovea. More non-congruent plumes appeared superior nasal to the fovea than in other quadrants. Of 129 plumes, 73 (56.6%) were associated with cRORA. Near cRORA, plumes were seen traveling in directions unrelated or opposite to the normal HFL (Supplementary Fig. 5). Accordingly, at the final visit (Supplementary Table 6), 48 of 129 (37.2%) plumes associated with cRORA were congruent with the local normal HFL and 25 of 129 (19.4%) were incongruent, with 16/129 (12.4%) of incongruent plumes located at the cRORA border. In contrast, all 6/129 (4.7%) plumes associated with iRORA were congruent with the HFL. Finally, 50/129 (38.8%) of plumes were not associated with either cRORA or iRORA.

### Laboratory study

To molecularly characterize normal and abnormal RPE cells, especially those corresponding to HRF, we used *ex vivo* OCT imaging to find human donor eyes with AMD (Fig. 4). HRF found in macular OCT B-scans were matched with pigmented cells in the subretinal space and retina of stained sections (Fig. 4). Atrophic AMD eyes with RPE plume in OCT exhibited pigmented cells in the outer nuclear layer (ONL) and HFL on histology (Fig. 4, C1 to C2 and F1 to F2). Results are based on 20 eyes of 20 donors (mean age 89.2 ± 5.0 years; 16 total AMD (7 early-intermediate, 6 atrophic, 3 neovascular, 4 control), with mean time from death to preservation of 3.9 ± 0.7 hours (Supplementary Table 7).

**Figure 4.**
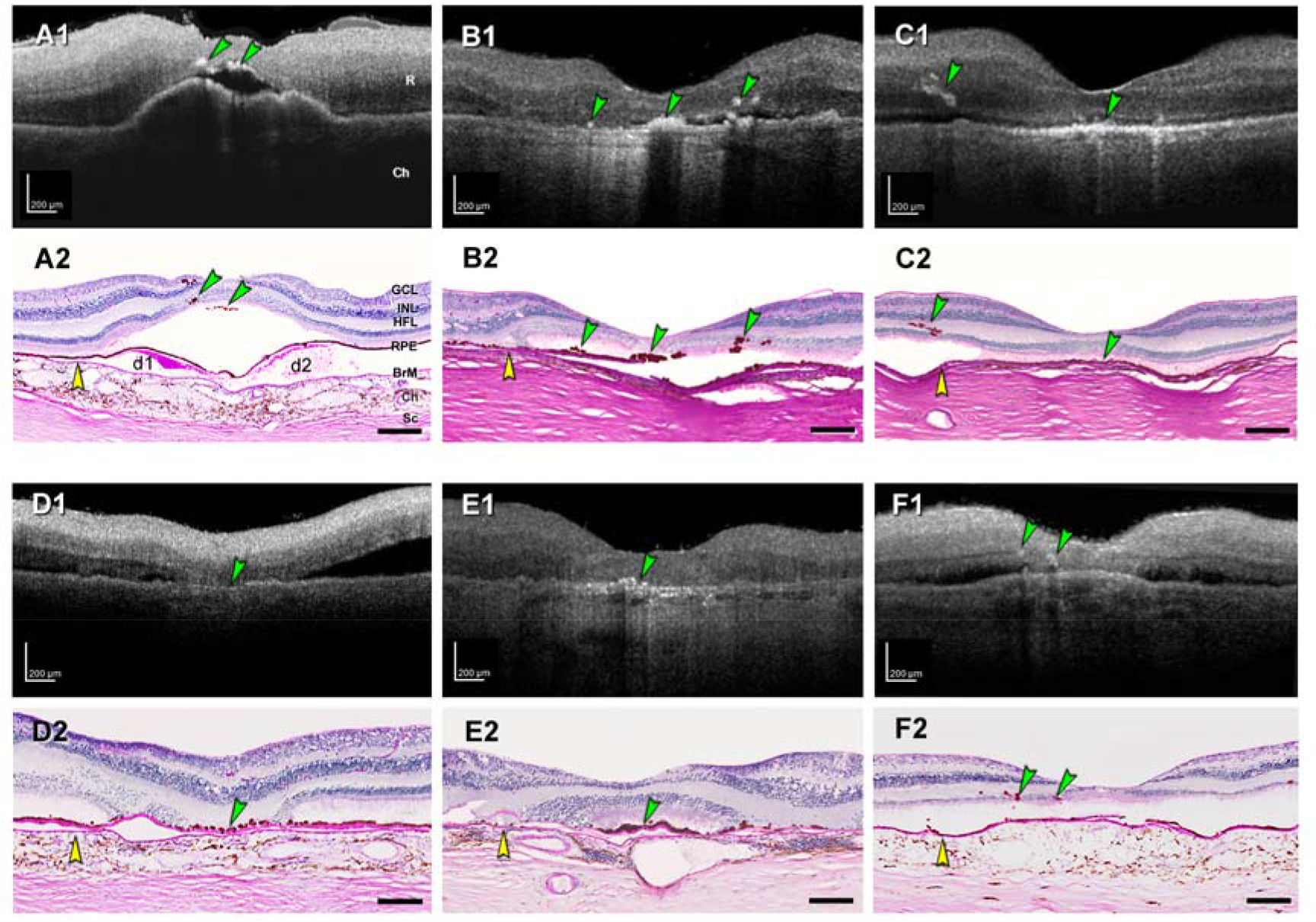
HRF in *ex vivo* OCT match intraretinal RPE on histology. Reflective features including hyperreflective foci (HRF) are visible in donor eyes (eye numbers in Supplementary Table 7) by *ex vivo* optical coherence tomography (A1 to F1). These were matched with RPE phenotypes in histology sections stained with periodic acid Schiff hematoxylin (PASH) (A2 to F2), to show Bruch’s membrane (BrM), basal laminar deposit, and drusen. In all panels, scale bars are 200 µm; yellow arrowheads indicate BrM, and green arrowheads indicate HRF in OCT and ectopic RPE in histology. (**A1** and **A2)** HRFs above soft drusen at fovea from early-intermediate AMD eye (#5). (**B1** and **B2)** HRFs at the fovea and nasal and temporal side, from atrophic AMD eye (#17). (**C1** and **C2)** RPE plume in atrophic AMD eye (#15). (**D1** and **D2**) Hyperreflective dots in OCT and dissociated RPE atop thick BLamD in the atrophic area. (**E1** and **E2**) Reflective dots under an island of surviving photoreceptors in the fovea center of an atrophic AMD eye (#12). (**F1** and **F2**) Multiple HRFs nasal to the fovea in an early-intermediate AMD eye (#7), corresponding to intraretinal RPE cells.

### Gain of immune function

We begin by demonstrating that cells of RPE origin label with antibodies against two well-known immune cell markers, Cluster of Differentiation (CD)68 and CD163. ^29^ CD68 is a transmembrane glycoprotein that is abundantly expressed by macrophages and widely used as a myeloid-specific marker. ^30^ CD68 is one member (LAMP-4) of a 5-protein family that together accounts for 50% of the protein in lysosome membranes. ^30^ In tissue sections (Fig. 5), positive controls for CD68 immunoreactivity are choroidal macrophages (Fig. 5D) and negative controls are healthy RPE (Fig. 5A). Fully pigmented cells in the neurosensory retina are strongly CD68+ (Fig. 5F). Importantly, individual sloughed cells in the subretinal space (Fig. 5C and 5F) are also positive, as are individual cells still within the layer (Fig. 5E). Dissociated RPE scattered in the atrophic area are universally labeled (Fig 5H and 5N), as are cells under the continuous layer (Fig. 5I), and within tubes of scrolled photoreceptors (Fig. 5J and 5K; outer retinal tubulation^31^). Some bilaminar (Fig. 5D) and melanotic (Fig. 5M) cells are CD68+.

**Figure 5.**
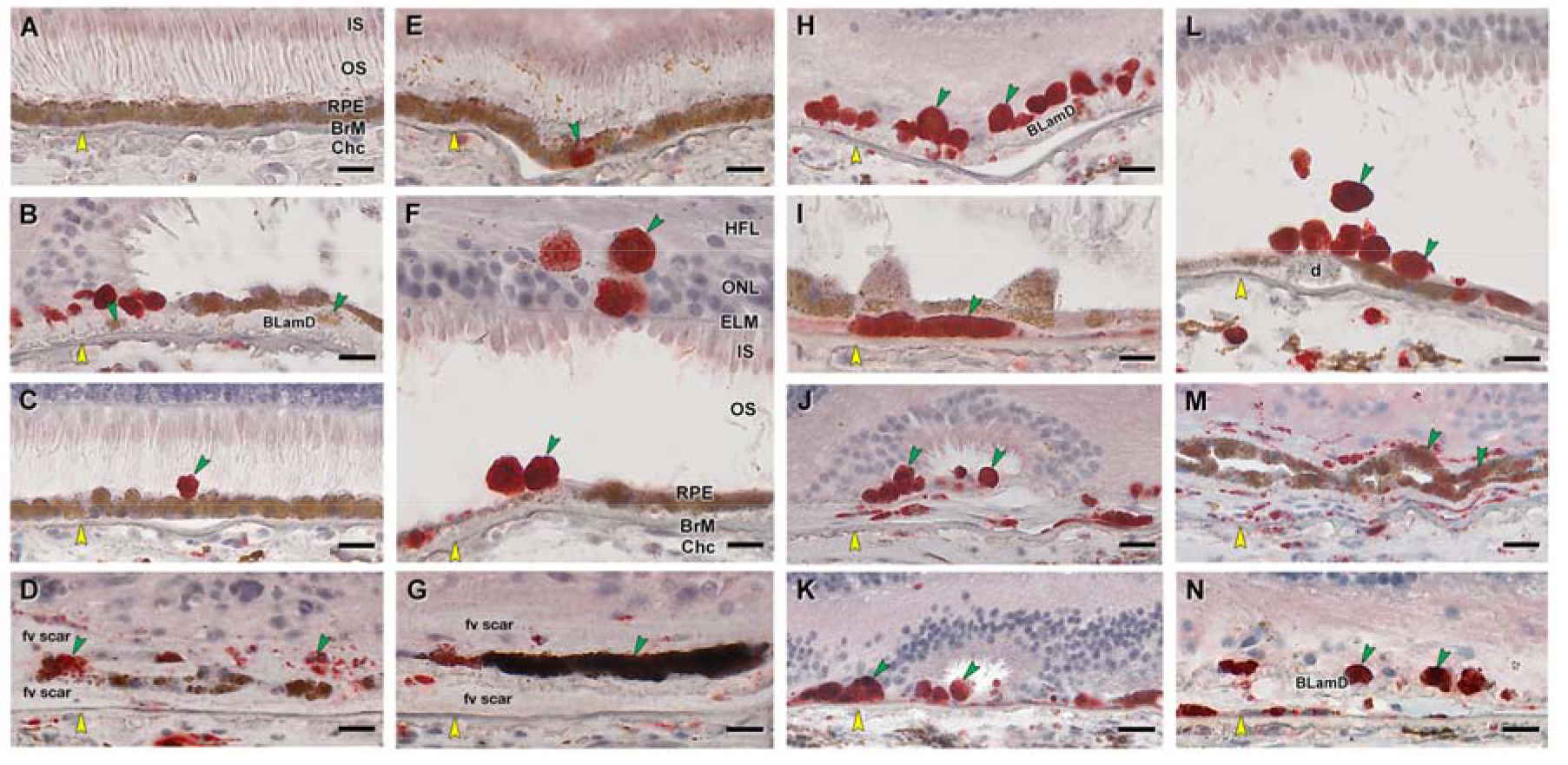
RPE corresponding to HRF and other abnormal cells are CD68+. Twenty donor eyes (16 AMD, 4 controls; eye numbers in Supplementary Table 7) were used for immunohistochemistry with mouse monoclonal anti-human CD68 (Supplementary Table 1) and red reaction product. Fourteen of 16 previously described morphologic phenotypes of RPE ^27^ were identified (Supplementary Table 2). All scale bars are 20 µm; yellow arrowheads indicate BrM, and green arrowheads indicate abnormal RPE phenotypes. (**A**) CD68- “Uniform” RPE, unremarkable (#1). (**B**) CD68-granule aggregates, released basally from ‘‘Shedding’’ RPE; atrophic AMD (#13). (**C**) CD68+ ‘‘Sloughed’’ RPE; atrophic AMD (#12). **D**. Both CD68+ and CD68- ‘‘Entombed’’ RPE; nvAMD (#18). (**C**) A single CD68+ “Nonuniform” RPE from a less-affected area, nvAMD (#20). (**F**) CD68+ ‘‘Intraretinal’’ RPE; early-intermediate AMD (#7). (**G**) Variable CD68 immunoreactivity in ‘‘Melanotic’’ RPE, nvAMD (#18). CD68 immunoreactivity is variable on heavily and light pigmented melanotic RPE. **H**. CD68+ “Severe” RPE; atrophic AMD (#13). (**I**) CD68+ ‘‘Subducted’’ RPE, atrophic AMD (#12). (**J**) CD68+ ‘‘Entubulated’’ RPE, atrophic AMD (#12). (**K)** CD68+ ‘‘Dissociated’’ RPE, atrophic AMD (#12). (**L**) CD68+ “Dissociated” RPE and “Sloughed” RPE, early-intermediate AMD (#7). (**M**) Variable CD68 immunoreactivity in ‘‘Bilaminar’’ RPE, nvAMD (#20). (**N**) CD68+ “Dissociated” RPE; atrophic AMD (#12).

CD163 is a high-affinity haptoglobin-hemoglobin scavenger found in bone marrow-derived macrophages ^32^ and in retinal cells resembling microglia. ^33-35^ The latter are phagocytes of embryologic yolk sac origin that reside in inner retina when quiescent and migrate into outer retina when photoreceptors are damaged. ^33^ Figure 6 shows that anti-CD163 labels inner retina cells but not the normal RPE layer, as expected. It does label abnormal RPE phenotypes in a pattern like that of CD68, with overall lighter reaction product. These include fully pigmented CD163+ cells in the ONL (Fig. 6Fand 6N), HFL (Fig. 6L), above (Fig. 6E and 6I) and below (Fig. 6I) the continuous RPE layer in the subretinal and sub-RPE-basal laminar spaces, respectively, and dissociated cells in atrophic areas (Fig. 6 H, 6J and 6K).

**Figure 6.**
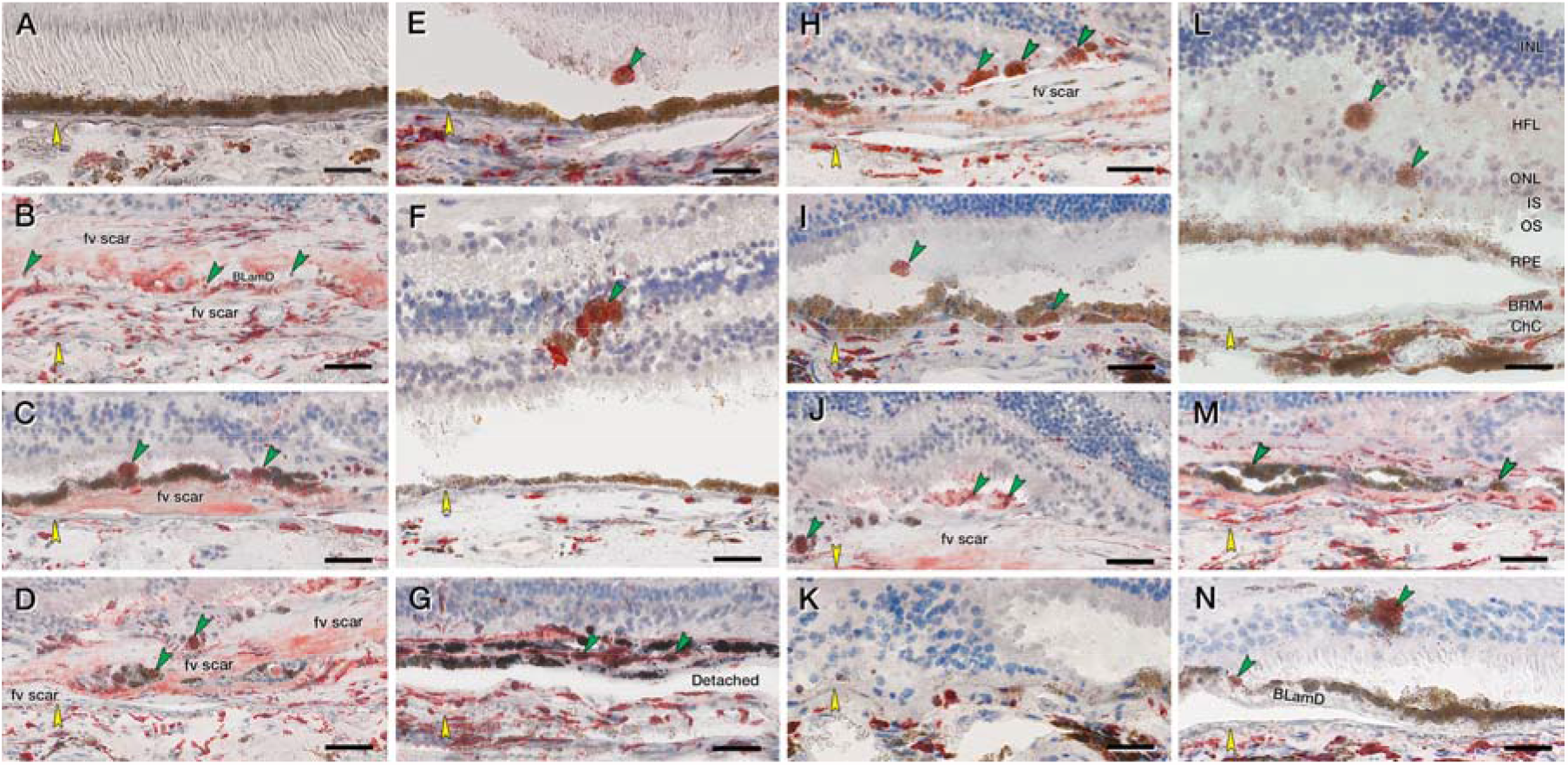
RPE corresponding to HRF and other abnormal cells are CD163+. Twenty donor eyes (16 AMD, 4 controls; eye number in Supplementary Table 7) were used for immunohistochemistry with mouse monoclonal anti-human CD163 (Supplementary Table 1) and red reaction product. Fourteen of 16 previously described morphologic phenotypes of RPE were identified (Supplementary Table 2). All scale bars are 20 µm; yellow arrowheads indicate BrM, and green arrowheads indicate abnormal RPE phenotypes. (**A**) CD163- “Uniform” RPE, unremarkable (#1). (**B**) CD163- ‘‘Shedding’’ RPE, nvAMD (#20). (**C**) CD163+ ‘‘Sloughed’’, nvAMD (#20). (**D**) Both CD163+ and CD163- are present in ‘‘Entombed’’ RPE, nvAMD (#20). (**E**) A single CD163+migrating RPE above “Nonuniform” RPE layer, atrophic AMD (#17). (**F**) CD163+ ‘‘Intraretinal’’ RPE, atrophic AMD (#15). (**G**) Variable CD163 immunoreactivities in ‘‘Melanotic’’ RPE, nvAMD (#20). (**H**) CD163+ “Severe” or “Very nonuniform” RPE, nvAMD (#20). (**I**) CD163+ ‘‘Subducted’’ and “Sloughed” RPE, atrophic AMD (#16). (**J**) CD163+ ‘‘Entubulated’’ and “Dissociated” RPE, nvAMD (#20). (**K**) There is no RPE in an area of ‘‘Atrophy without BLamD’’, atrophic AMD (#16). (**L**) CD163+ “Dissociated” and “Intraretinal” RPE, atrophic AMD (#16). (**M**) Variable CD163 immunoreactivity in ‘‘Bilaminar’’ RPE, nAMD (#20). (**N**) CD163+ “Intraretinal” RPE at ‘‘Atrophy with BLamD’’ area, atrophic AMD (#14).

### Loss of retinoid function

Next, we show that abnormal RPE cells cease to exhibit detectable quantities of retinoid processing proteins essential for vision. Rod-mediated (dim light) vision is affected early in AMD, because rods depend on the choriocapillaris-Bruch’s membrane-RPE complex. ^36^ Cone-mediated (bright light) vision, which is also sustained by Müller glia and retinal vessels, ^37^ persists into later disease.

We begin with RPE65, the isomerohydrolase of the classic visual cycle. RPE65 catalyzes the conversion of retinyl esters to 11-cis-retinol, which is subsequently oxidized to 11-cis-retinal, required for visual transduction by rod and cone photoreceptors. ^38^ Figure 7 shows that in AMD eyes, intraretinal RPE in ONL (Fig. 7F) and individual sloughed cells overlying the continuous layer (Fig. 7C and 7L) lack RPE65-immunoreactivity. Cells in areas of photoreceptor degeneration and depletion are also RPE65-, including dissociated (Fig. 7N), subducted (Fig. 7I) and entubulated (Fig. 7J) cells. Importantly, some individual cells in the continuous layer are RPE65-(Fig. 7F and 7H). Interestingly some bilaminar cells within fibrotic scars of neovascular AMD remain RPE65+ (Fig. 7M).

**Figure 7.**
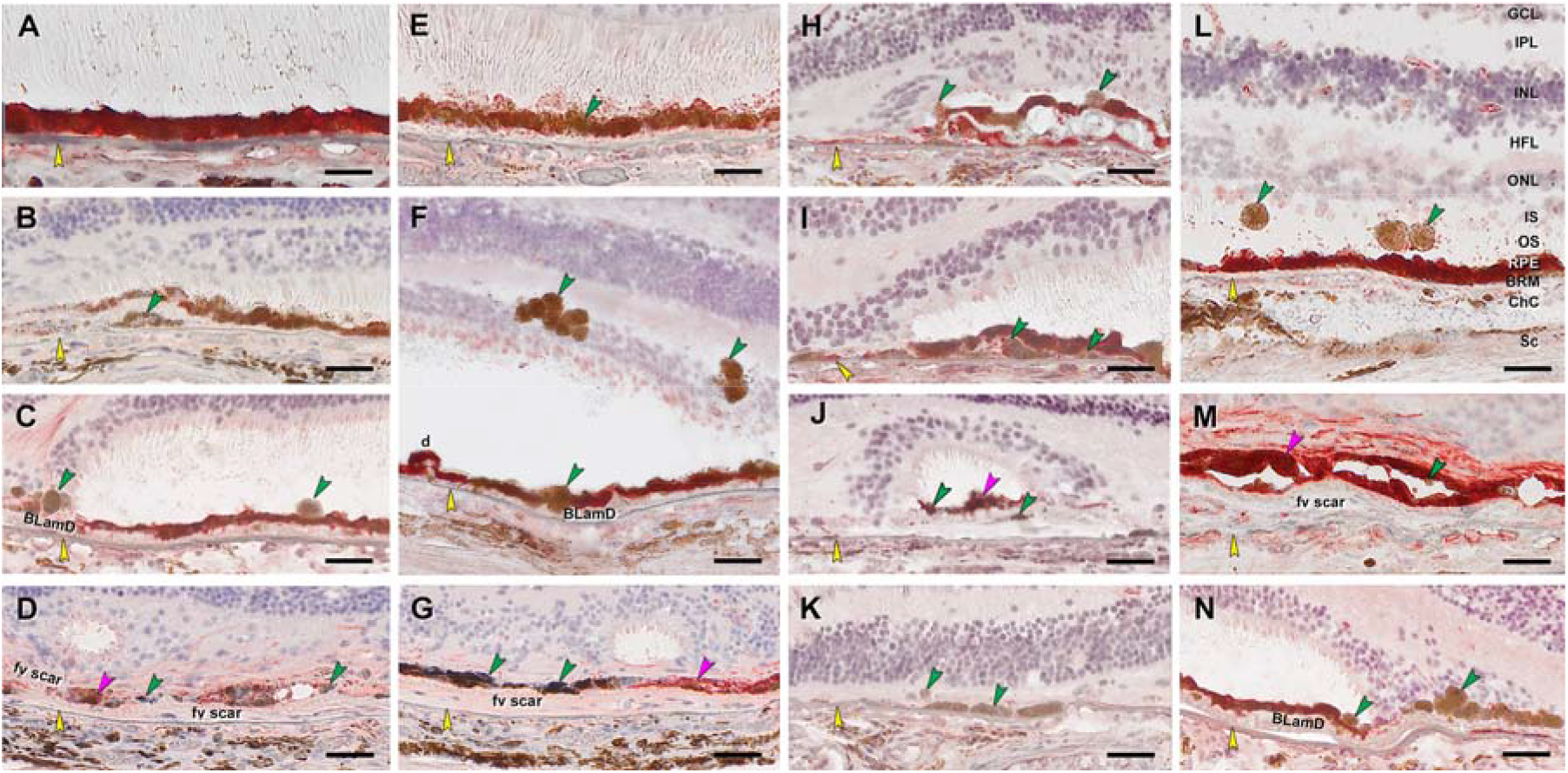
RPE corresponding to HRF are RPE65-, as are other abnormal cells. Twenty donor eyes (16 AMD, 4 controls; eye numbers in Supplementary Table 7) were used for immunohistochemistry with mouse monoclonal anti-human RPE65 (Supplementary Table 1) and red reaction product. Fourteen of 16 previously described morphologic phenotypes of RPE ^27^ were identified (Supplementary Table 2). All scale bars are 20 µm; yellow arrowheads indicate BrM; green and fuchsia arrowheads indicate RPE65- and RPE65+ abnormal RPE phenotypes, respectively. (**A**) RPE65+ “Uniform” RPE, unremarkable (#1). (**B**) RPE65- ‘‘Shedding’’ RPE, nvAMD (#20). (**C**) RPE65- ‘‘Sloughed’’ RPE, atrophic AMD (#13). (**D**) Both RPE65+ and REP65- ‘‘Entombed’’ RPE, nvAMD (#18). (**E**) A few RPE65- “Nonuniform” RPE, early-intermediate AMD (#6). (**F**) RPE65- ‘‘Intraretinal’’ RPE, early-intermediate AMD (#11). (**G**) Variable RPE65 immunoreactive ‘‘Melanotic’’ RPE, nvAMD (#18). (**H**) RPE65- “Severe” RPE, atrophic AMD (#12). (**I**) RPE65- ‘‘Subducted’’ RPE, atrophic AMD (#12). (**J**) Both RPE65+ and RPE65- ‘‘Entubulated’’ RPE, atrophic AMD (#12). (**K**) RPE65-RPE at ‘‘Atrophy without BLamD’’ area, atrophic AMD (#12). (**L**) RPE65- “Dissociated” RPE, atrophic AMD (#16). (**M**) Both RPE65- and RPE65+ ‘‘Bilaminar’’ RPE, nvAMD (#20). (**N**) RPE65-RPE at ‘‘Atrophy with BLamD’’ area, atrophic AMD (#13).

Cellular retinal binding protein (CRALBP) is important in the RPE-based visual cycle. It is also expressed by Müller glia in a second visual cycle thought to supply cones. ^37, 39^ CRALBP is a 36-kDa water-soluble protein with 2 conformational states facilitating intracellular transport of hydrophobic 11-cis retinoids. ^40^ Figure 8 shows that healthy RPE has a strong CRALBP signal (Fig. 8A). In AMD, ectopic RPE in the ONL, recognizable by pigment load, are CRALBP-(Fig. 8F and 8L). Individual sloughed cells (Fig. 8H and 8L) and in-layer cells (Fig. 8E) are also negative as are pigmented cells in atrophic areas (dissociated, Fig. 8C, 8K and 8N; subducted, Fig. 8I). Shed granule aggregates ^15, 41^ are CRALBP-, under a continuous CRALBP+ layer (Fig. 8B, J).

**Figure 8.**
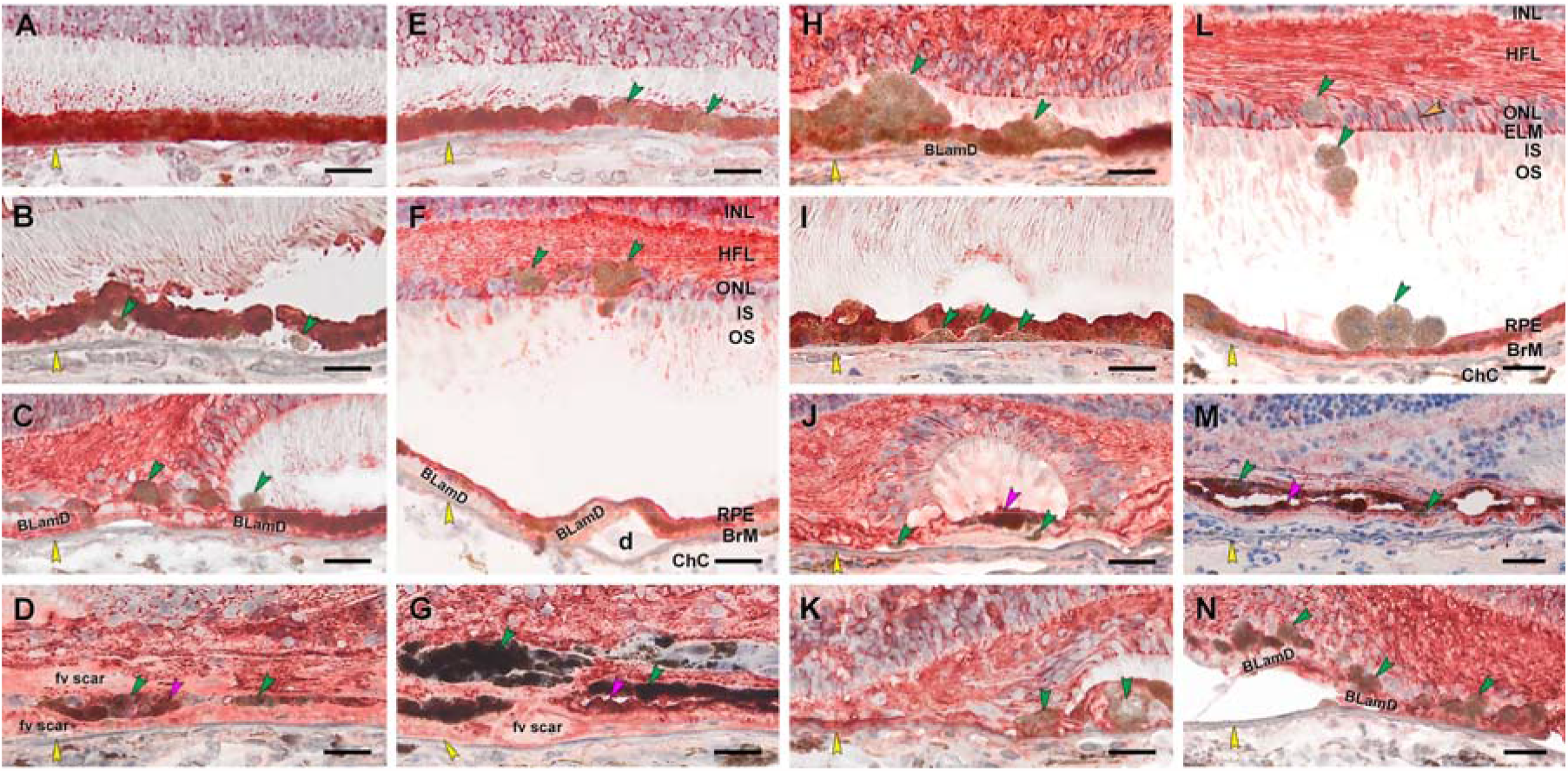
Abnormal RPE is CRALBP-, Müller glia are CRALBP+. Twenty donor eyes (16 AMD, 4 controls; eye numbers in Supplementary Table 7) were used for immunohistochemistry with mouse monoclonal anti-human CRALBP (Supplementary Table 1) and red reaction product. Fourteen of 16 previously described morphologic phenotypes of RPE ^27^ were identified (Supplementary Table 2). All scale bars are 20 µm. Yellow arrowheads indicate BrM. Green and fuchsia arrowheads indicate CRALBP- and CRALBP+ abnormal RPE phenotypes, respectively. In panel L, labeled Müller glia are seen at the external limiting membrane (ELM), pericellular baskets in ONL (orange arrowhead), and parallel fibers in HFL. (**A**) CRALBP+ “Uniform” RPE, unremarkable (#1). (**B**) CRALBP- ‘‘Shedding’’ RPE, early-intermediate AMD (#6). (**C**) CRALBP- ‘‘Sloughed’’ RPE, atrophic AMD (#13). (**D**) Both CRALBP+ and CRALBP- ‘‘Entombed’’ RPE nvAMD (#18). (**E**) CRALBP- “Nonuniform” RPE nvAMD (#18). (**F**) CRALBP- ‘‘Intraretinal’’ RPE, early-intermediate AMD eye (#7). (**G**) Both CRALBP+ and CRALBP- ‘‘Melanotic’’ RPE, nvAMD (#18). (**H**) CRALBP- “Severe” RPE, atrophic AMD (#12). (**I**) CRALBP- ‘‘Subducted’’ RPE, atrophic AMD (#12). (**J**) Both CRALBP+ and CRALBP- ‘‘Entubulated’’ RPE, atrophic AMD (#12). (**K**) CRALBP-RPE at ‘‘Atrophy without BLamD’’ area, atrophic AMD (#12). (**L**) CRALBP- “Dissociated” RPE, early-intermediate AMD (#7). (**M**) Variable CRALBP immunoreactivity in ‘‘Bilaminar’’ RPE, nvAMD (#20). (**N**) Variable CRALBP+ and CRALBP-RPE at ‘‘Atrophy with BLamD’’ area, nvAMD (#7).

In contrast to RPE, Müller glia remain strongly CRALBP+ in their cross-retinal course, regardless of the degree of photoreceptor loss. Distinctive features (Fig. 8L) are the external limiting membrane (junctional complexes between glia and photoreceptors) ^42^ and pericellular baskets ^43^ around ONL cell bodies. In the HFL of areas with lesser disease involvement, stained Müller glia are parallel and straight (ordered, ^7^ Fig. 8L). At the border of atrophy (descent of the external limiting membrane, ^44^ Fig. 8C, 8K and 8N) and outer retinal tubulation (Fig. 8J), CRALBP+ fibers curve down towards Bruch’s membrane and then head laterally, parallel to it. In some locations immunolabeled fibers zig-zag in parallel, presenting cross-sections of dots and lines (disordered, ^7^ Fig. 8F and 8N). CRALBP signal also appears in fibrotic scars resulting from neovascular AMD (Fig. 8D, 8G and 8M).

### Semi-quantitative assessment of immunohistochemistry

The number of stained cells in each RPE phenotype was assessed a semi-quantitatively (0, not stained; 1, some cells of a phenotype stained; 2, all cells stained). These scores were summed and expressed as a percent of the highest possible score, as if all phenotypes had been stained completely by each antibody. Supplementary Figure 6A shows that all RPE phenotypes were found in immuno-stained sections of these 16 AMD eyes, with similar patterns in neighboring sections. Supplementary Figure 6B graphically summarizes the marker studies. Age-normal uniform and non-uniform RPE is positive for retinoid markers and negative for immune markers. Abnormal RPE phenotypes have overall less retinoid immunoreactivity (although some stained cells were found at 5%) and universally more immune marker immunoreactivity (up to 75%). Thus, cells corresponding to HRF in OCT are part of an overall pattern of transdifferentiation resulting in gain-of-function and loss-of-function of RPE.

### Histology of plume lifecycle

Our clinical study indicated that RPE plumes have lifecycles that are observable in vivo. We previously showed in type 3 (intraretinal) neovascularization that RPE cells contact the deep capillary plexus. ^18^ Figure 9, A to C accordingly shows RPE plume cells crossing the HFL to approach and encircle capillaries of this plexus. Cells are nucleated and contain numerous RPE-characteristic spindle-shaped melanosomes and melanolipofuscin granules. Fine processes extend along a capillary (Fig. 9D).

**Figure 9.**
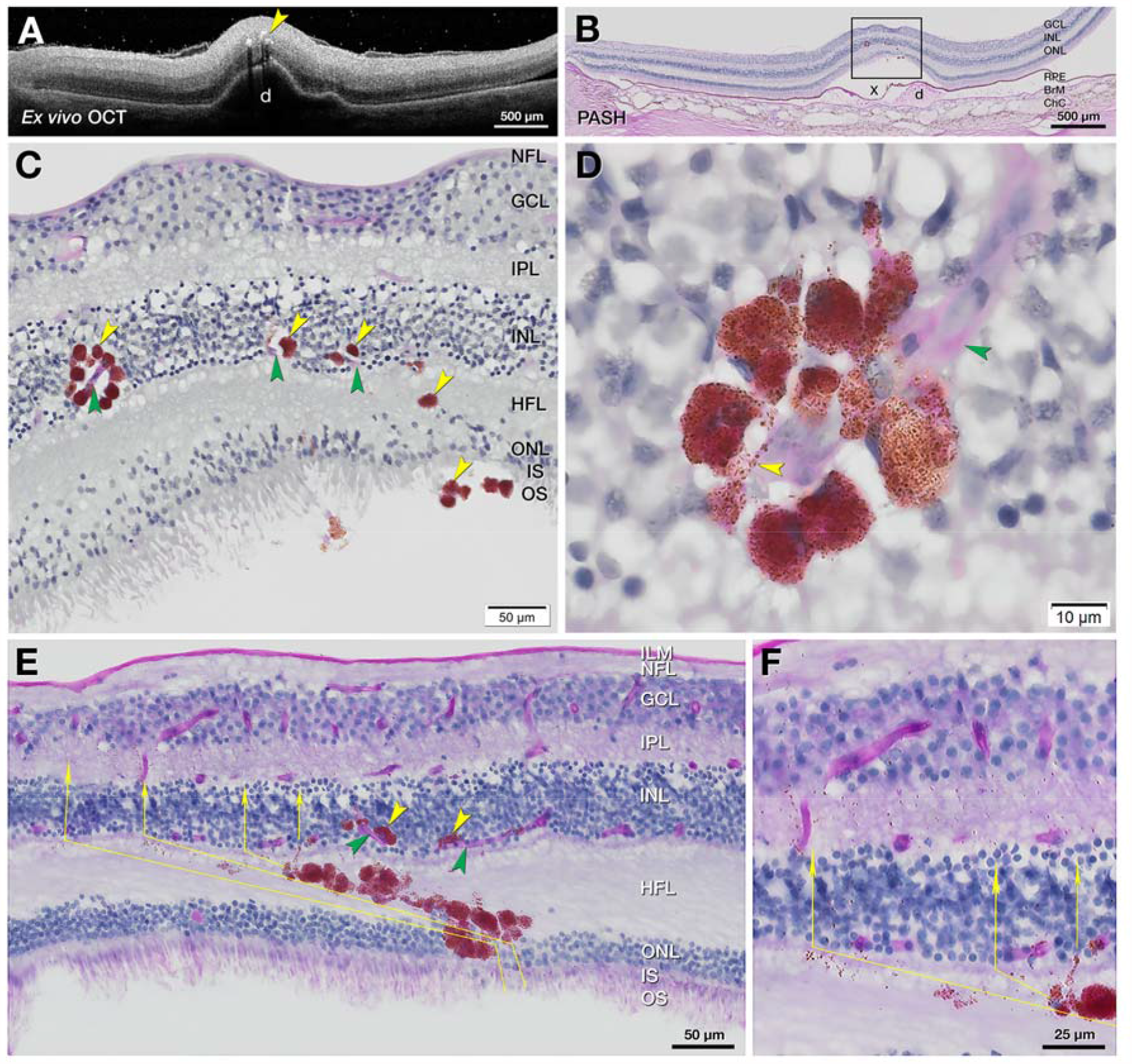
RPE migrate toward vessels and follow Müller glial columns. One eye from each of two donors aged 86-90 years (#5 and #15, Supplementary Table 7) were subjected to *ex vivo* OCT images in A (Fig. 4, C1 and C2) and PASH staining with cryosections in B, E and F. (**A**) *Ex vivo* OCT of donor eye (#5) shows HRF (yellow arrowhead) over a large druse. (**B**) Corresponding PASH histologic section 290 µm superior to the fovea with a big druse (d), artifactually detached (x) from the RPE, and thinner ONL. (**C**) Magnified RPE rosette in B (yellow arrowheads) crossing the HFL and approaching retinal capillaries (green arrowheads). (**D**) Nucleated RPE rosette from panel C (adjacent section). RPE-characteristic spindle-shaped melanosomes and melanolipofuscin impart a brown hue. The surrounded capillary is stained light pink (green arrowhead) with endothelial cell nuclei. Processes containing RPE organelles extend along the capillaries. (**E** and **F**) In another plume, RPE cells and detritus take a Z-shaped course. Fully pigmented RPE cells cross the obliquely oriented fibers of the HFL, with individual and grouped organelles in the lead (thin yellow arrows). The path turns vertically to enter and cross the INL and IPL.

We previously showed RPE organelles in the HFL of photoreceptor-depleted atrophic areas, ^7^ suggesting that Müller glia ingested and cleared debris from degenerating RPE. Here we show region-specific distribution of RPE organelles, consistent with the local trajectory of Müller glia. In the macula, Müller glia are Z-shaped. ^45^ Fibers are oriented vertically in the ONL, obliquely in the HFL, and vertically in the OPL and inner retinal layers (Supplementary Fig. 1). In plumes near the fovea (Fig. 9E and 9F; Fig. 6N), fully pigmented RPE cells obliquely cross the HFL, with individual and grouped organelles in the lead. The path turns vertically at the INL and IPL, following a Z-shaped course. In contrast, in the peripheral retina, where Müller glia are vertical, Supplementary Figure 7 shows melanosomes on a vertical track away from RPE above a druse.

## DISCUSSION

For RPE plume, one HRF type in AMD, we established a clinical lifecycle, including both cRORA and resolution without cRORA, with evidence of clearance by Müller glia. HRF are one of four OCT indicators in a composite risk score for AMD progression (with soft and calcified drusen, plus subretinal drusenoid deposit). ^46^ Of these four, HRF confer the largest risk (odds ratio at 24 months, 5.21 (3.29-8.26)),^13^ yet have received histologic investigation only recently, a gap that is herein addressed. We provide strong evidence that ectopic RPE can manifest as HRF, without excluding the possibility that other cells, such as microglia, macrophages, or both, may also correspond to HRF, as they do in nvAMD. ^18, 20^ We further show that sloughed and intraretinal cells corresponding to HRF are two of many abnormal RPE phenotypes undergoing molecular transdifferentiation.

Our parallel findings of CD68 and CD163 immunoreactivity in all abnormal RPE phenotypes adds important new information to a decades-old debate ^47, 48^ about whether RPE, a phagocyte itself, shares or assumes properties of other phagocytes. This investigation like our previous studies used high resolution comprehensive microscopy to link cells with characteristic RPE organelles ^15, 16^ to a time course of activity visible through longitudinal clinical imaging. Our cellular phenotyping system ^15^ built on prior literature and is used by others. ^49, 50^ We posited that these phenotypes represented discrete steps towards death, migration, or transdifferentiation of individual cells that could be followed over time in OCT due to high RPE reflectivity. In prior electron microscopic analysis of pigmented cells corresponding to HRF ^51^ we did not observe phagolysosomes, i.e., cytoplasmic vacuoles formed by phagosomes with ingested material fusing with lysosomes, as expected from phagocyte activity. In a separate study of HRF over a large drusenoid pigment detachment, ^17^ all detectable HRF could be attributed to fully pigmented and nucleated intraretinal cells. In clinical cases of that prior study, ^17^ HRF appeared in the retina vertically above and several months after disturbances of the RPE layer. Our current data further show that individual in-layer cells and all abnormal phenotypes exhibit CD68 and CD163 immunoreactivity. We cannot exclude the possibility that phagocytes ingest and retain characteristic RPE organelles, seamlessly intercalate into the layer, and either become abnormal phenotypes or induce them. However, extensive transdifferentiation in response to common perturbations is also a parsimonious explanation.

We emphasize that snapshot immuno-detection of CD68, a lysosomal protein of unknown function reliably found in macrophages of myeloid origin, ^30^ does not indicate that labeled cells have assumed macrophage behavior. Our data do not imply that macrophages of hematopoietic origin previously shown in choroid and sub-RPE-BL space ^52, 53^ started as RPE. Our data do not bear on the identity of CD68+ cells seen by others, e.g., high-passage cultured RPE ^54^ and surgically excised neovascular membranes. ^55, 56^ In rapidly preserved eyes with experimental laser-injury, individual detached cells atop an intact RPE layer are CD68+ ^57^(compare to Fig. 5C and Fig. 6C). Recent authors interpreted CD68+ cells in aqueous after RPE tear ^58^ and in proliferative vitreoretinopathy ^59^ as RPE. CD68+ cells in the outer junctional zone of atrophy in AMD were demonstrated by laser confocal microscopy and immunofluorescence. ^49^ This technique does not directly reveal melanin, and the authors did not comment on cellular identity. ^49^

Our data illuminate subducted cells on the inner collagenous layer of Bruch’s membrane containing RPE organelles and originally called “spent RPE”. ^60^ Using single continuous high-resolution histologic sections of eyes with cRORA, we saw plausible transitional morphologies between fully pigmented RPE outside atrophic areas and flattened cells with fewer organelles in the sub-RPE-BL space. ^16^ We hypothesized that dissociated RPE in an atrophic area, ^61^ loosed from constraining junctional complexes, dove under the intact layer and moved into the outer junctional zone to participate in atrophy expansion (Fig. 1). ^16^ Herein we show CD68+ and CD163+ cells and highly pigmented RPE65- and CRALBP-cells in the same sub-RPE-BL tissue compartment of the same eyes. The sub-RPE-BL space can harbor macrophages of myeloid origin, multinucleate giant cells, and fibroblasts. ^62^ It is possible that co-localized CCR2 and CD18 immunoreactivity in these locations, attributed to macrophages, ^63^ may represent subducted and dissociated RPE, respectively. Multi-label immunohistochemistry is needed to confirm how many subducted cells are RPE-derived. Macrophages that co-express CD68 and CCR7 can be assigned to a ‘‘proinflammatory’’ M1 subset, whereas CD68+ CD163+ macrophages most likely belong to an ‘‘anti-inflammatory’’ M2 subset. ^64^ Longitudinal studies with higher resolution OCT ^65, 66^ may make it possible to detect subducted cells and determine if they promote or retard atrophy expansion, of value to validating this regulatorily-approved anatomic endpoint.

Our expectation of plume congruence with HFL trajectory was largely met and of note was non-congruence near cRORA (Supplementary Table 6). This is likely due to subsidence (sinking) of the OPL, HFL, and ONL, a committed step toward cRORA onset. ^67^ Subsidence is visible where photoreceptors focally shorten and are scrolled by Müller glia. ^7, 68^ Our data on plume trajectories and CRALBP immunohistochemistry thus add to prodigious activities of Müller glia in AMD. These include descent and curving of the ELM (border of atrophy), reactive gliosis, adherent glial seal in areas of photoreceptor depletion, and invasion of individual drusen. ^31, 44, 69, 70^ CRALBP immunohistochemistry confirmed HFL fiber geometry as well as retinoid capacity of Müller glia, still supported by retinal circulation late in disease. Single and grouped RPE organelles paralleling Müller glia ^7, 71^ are presumably carried intracellularly. In contrast RPE cell bodies travel between HFL fibers (Fig. 8L and Fig. 9) following a trail of organelle detritus. Active glial disassembly and removal of fragments is suspected. Because almost 40% of plumes were not associated with atrophy at follow-up (Supplementary Table 6), plumes should be considered indicators not predictors. Factors underlying variable plume outcome in our study include inclusion of patients with established disease, short observation period, and a “come and go” behavior consistent with cells subject to constant pro-migratory stimuli being dismantled *en route*.

Given evidence that many RPE cells are transdifferentiating, HRF may indicate risk by revealing an overall level and type of disease activity. Epithelial-mesenchyme-transition (EMT) is one candidate activity. In this complex process, cells lose apical-basal polarity, modulate cytoskeleton, and reduce cell-cell adhesion. ^72^ A defining feature of an epithelium is localization within organs stabilized by a basal lamina and integrin-mediated interactions with extracellular matrix. Epithelial cells that detach from extracellular matrix normally undergo a programmed cell death called anoikis. ^73, 74^ Tumor cells of epithelioid origin can escape anoikis by EMT. Thus, they assume amoeboid morphology, become motile and invasive, and travel in the circulation to other organs, with most dying *en route*. EMT of RPE has been suspected in AMD due to findings of characteristic molecular markers (vimentin ^76^ and the transcription factor Snail1 ^21^) and the presence of HRF. ^75^ Another contributor is the separation of the RPE-BL from its native basal lamina by BLamD, ^23, 77^ an extracellular matrix that also contains other components. RPE cells undergoing EMT might indeed resist cell death ^21^ but they may also be dysfunctional, due to gain-and loss-of-function suggested by our data. One factor promoting EMT over anoikis is hypoxia, ^74^ of high AMD relevance due to age- and disease-related loss of choroid and choriocapillaris ^78-81^ and the distancing of RPE from choriocapillaris by BLamD and drusen. ^82^ Thus we showed RPE cells not only contacting the deep capillary plexus (**Fig. 9** ^17, 18^) but also aiming towards and clustering around vessels as a seemingly coordinated group (**Fig. 9**) rather than as single cells. Parallels with cancer are appropriate due to the unusual metabolic demands of photoreceptors and their supporting system compared to other neurons. The retina, of all body tissues, shares with tumors a dependence on aerobic glycolysis (the Warburg effect). ^83-85^ A consensus group of cancer researchers recently advocated monitoring cellular properties as part of the main criteria for EMT, along with molecular markers. ^72^ The high OCT visibility of RPE makes it uniquely possible to view these cellular activities in AMD.

Strengths of the clinical study are high-quality images and long follow-up on known progression risk factors. Limitations include the small number of patients and lack of independently assessed HFL trajectory with directional OCT. ^86^ Strengths of the laboratory study are short post-mortem specimens with attached retinas, comprehensive photomicroscopy to visualize pigment, a color-contrasting readout for immunoreactivity, and a cross-sectional format conducive to OCT comparison. Limitations addressable in future studies include lack of clinical records with pre-mortem imaging, lack of correlative data from flat mounts to show early RPE pathology, limited assessments of RPE adjacent to AMD deposits, and lack of double-label experiments to determine cell-by-cell overlap among the four markers. Nevertheless, this is most detailed characterization of RPE pathology in AMD to date in either clinical or laboratory settings. HRF are a consensus OCT feature in AMD, ^10^ encouraging longitudinal studies in other clinical centers, ideally with comprehensive *en face* OCT and fundus autofluorescence imaging to determine the proportion of HRF that are RPE-derived. Our data can propel new biomarker and therapeutic strategies for a common and currently untreatable cause of impaired vision.

## Data Availability

Available on request.

## Abbreviations

AMD: Age-related macular degeneration
OCT: optical coherence tomography
HRF: hyperreflective foci
BLamD: basal laminar deposit
BrM: Bruch’s membrane
Ch: choroid
ChC: choriocapillaris
cRORA: complete RPE and outer retinal atrophy
d: drusen
ELM: external limiting membrane
EZ: ellipsoid zone
fv scar: fibrovascular scar
GCL: Ganglion cell layer
HFL: Henle fiber layer
ILM: internal limiting membrane
IPL: inner plexiform layer
INL: inner nuclear layer
IS: inner segments of photoreceptors
iRORA: complete RPE and outer retinal atrophy
IZ: interdigitation zone
nvAMD: neovascular AMD
ONL: outer nuclear layer
OPL: outer plexiform layer
OS: outer segments of photoreceptors
R: retina
RPE: retinal pigment epithelium
Sc: sclera
SRS: subretinal space
x: detachment

## SUPPLEMENTARY MATERIALS

### Supplementary Figures

Supplementary Figure 1. Henle fibers in the human macula.

Supplementary Figure 2. Positive and negative controls for immunohistochemistry

Supplementary Figure 3. Retinal pigment epithelium plumes exhibit directionality

Supplementary Figure 4. Duration and evolution of plumes

Supplementary Figure 5. Variations in RPE plume trajectory in atrophy.

Supplementary Figure 6. Gain- and loss-of-function of RPE and RPE-derived cells

Supplementary Figure 7. Dispersing melanosomes follow regionally specific trajectories of Müller glia

### Supplementary Tables

Supplementary Table 1. Antibodies used

Supplementary Table 2. Morphologic phenotypes of RPE and its basal lamina in AMD

Supplementary Table 3. Demographics of clinic patients and study eyes

Supplementary Table 4. Plume disposition at final visit

Supplementary Table 5. Plume trajectory relative to HFL and macular quadrant

Supplementary Table 6. Plume association with atrophy

Supplementary Table 7. Characteristics of donor eyes

## Acknowledgments

We thank Advancing Sight Network (formerly the Alabama Eye Bank) for timely retrieval of donor eyes and David Fisher for graphics in Figure 1.

## Author contributions

C.A.C. designed, supervised study and wrote the manuscript. D.C. designed and performed immunohistochemistry experiments. B.L. designed and conducted clinical studies and wrote the manuscript. J.M. conducted *ex vivo* imaging, tissue cryosections, and microscopy. D.K. made Fig. S1. T.A. edited manuscript. L.A.Y. and K.B.F designed and supervised clinical study. K.B.F. edited manuscript. C.A.C. and T.A. raised funding.

## Data and materials availability

All other data associated with this study are present in the paper or the Supplementary Materials. Materials are available upon request from the corresponding author.

## References

1. Flaxman SR, Bourne RRA, Resnikoff S, et al. Global causes of blindness and distance vision impairment 1990-2020: a systematic review and meta-analysis. The Lancet Global Health 2017;5:e1221–e1234. PMID 29032195

2. Maguire MG, Martin DF, Ying GS, et al. Five-year outcomes with anti-vascular endothelial growth factor treatment of neovascular age-related macular degeneration: the Comparison of Age-Related Macular Degeneration Treatments Trials. Ophthalmology 2016;123:1751–1761. PMID 27156698

3. Chen L, Messinger JD, Kar D, Duncan JL, Curcio CA. Biometrics, impact, and significance of basal linear deposit and subretinal drusenoid deposit in age-related macular degeneration. Invest Ophthalmol Vis Sci 2021;62:33. PMID 33512402

4. Keane PA, Patel PJ, Liakopoulos S, Heussen FM, Sadda SR, Tufail A. Evaluation of age-related macular degeneration with optical coherence tomography. Surv Ophthalmol 2012;57:389–414. PMID 22898648

5. Guymer RH, Rosenfeld PJ, Curcio CA, et al. Incomplete retinal pigment epithelial and outer retinal atrophy (iRORA) in age-related macular degeneration: CAM Report 4. Ophthalmology 2020;127:394–409. PMID 31708275

6. Pollreisz A, Neschi M, Sloan KR, et al. An atlas of human retinal pigment epithelium organelles significant for clinical imaging. Invest Ophthalmol Vis Sci 2020;61:13. PMID 32648890

7. Li M, Huisingh C, Messinger JD, et al. Histology of geographic atrophy secondary to age-related macular degeneration: a multilayer approach. Retina 2018;38:1937–1953. PMID 29746415

8. Holz FG, Sadda S, Staurenghi G, et al. Imaging protocols for clinical studies in age-related macular degeneration – recommendations from Classification of Atrophy (CAM) Consensus Meeting. Ophthalmology 2017;124:464–478. PMID 28109563

9. Sadda SR, Guymer R, Holz FG, et al. Consensus definition for atrophy associated with age-related macular degeneration on optical coherence tomography: CAM Report 3. Ophthalmology 2018;125:537–548. PMID 29103793

10. Jaffe GJ, Chakravarthy U, Freund KB, et al. Imaging features associated with progression to geographic atrophy in age-related macular degeneration: CAM Report 5. Ophthalmology Retina 2020;12/20/20 online. PMID 33348085

11. Ouyang Y, Heussen FM, Hariri A, Keane PA, Sadda SR. Optical coherence tomography-based observation of the natural history of drusenoid lesion in eyes with dry age-related macular degeneration. Ophthalmology 2013;120:2656–2665. PMID 23830761

12. Christenbury JG, Folgar FA, O’Connell RV, Chiu SJ, Farsiu S, Toth CA. Progression of intermediate age-related macular degeneration with proliferation and inner retinal migration of hyperreflective foci. Ophthalmology 2013;120:1038–1045. PMID 23352193

13. Nassisi M, Lei J, Abdelfattah NS, et al. OCT risk factors for development of late age-related macular degeneration in the fellow eyes of patients enrolled in the HARBOR study. Ophthalmology 2019;126:1667–1674. PMID 31281056

14. Waldstein SM, Vogl W-D, Bogunovic H, Sadeghipour A, Riedl S, Schmidt-Erfurth U. Characterization of drusen and hyperreflective foci as biomarkers for disease progression in age-related macular degeneration using artificial intelligence in optical coherence tomography. JAMA Ophthalmology 2020;138:740–747. PMID 32379287

15. Zanzottera EC, Messinger JD, Ach T, Smith RT, Freund KB, Curcio CA. The Project MACULA retinal pigment epithelium grading system for histology and optical coherence tomography in age-related macular degeneration. Invest Ophthalmol Vis Sci 2015;56:3253–3268. PMID 25813989

16. Zanzottera EC, Messinger JD, Ach T, Smith RT, Curcio CA. Subducted and Melanotic cells in advanced age-related macular degeneration are derived from retinal pigment epithelium. Invest Ophthalmol Vis Sci 2015;56:3269–3278. PMID 26024109

17. Balaratnasingam C, Messinger JD, Sloan KR, Yannuzzi LA, Freund KB, Curcio CA. Histologic and optical coherence tomographic correlations in drusenoid pigment epithelium detachment in age-related macular degeneration. Ophthalmology 2017;124:644–656. PMID 28153442

18. Li M, Dolz-Marco R, Messinger JD, et al. Clinicopathologic correlation of anti-vascular endothelial growth factor-treated type 3 neovascularization in age-related macular degeneration. Ophthalmology 2018;125:276–287. PMID 28964579

19. Jopling C, Boue S, Izpisua Belmonte JC. Dedifferentiation, transdifferentiation and reprogramming: three routes to regeneration. Nat Rev Mol Cell Biol 2011;12:79–89. PMID 21252997

20. Pang C, Messinger JD, Zanzottera EC, Freund KB, Curcio CA. The Onion Sign in neovascular age-related macular degeneration represents cholesterol crystals. Ophthalmology 2015;122:2316–2326. PMID 26298717

21. Ghosh S, Shang P, Terasaki H, et al. A role for betaA3/A1-crystallin in Type 2 EMT of RPE cells occurring in dry age-related macular degeneration. Invest Ophthalmol Vis Sci 2018;59:AMD104–AMD113. PMID 30098172

22. Staurenghi G, Sadda S, Chakravarthy U, Spaide RF. Proposed lexicon for anatomic landmarks in normal posterior segment spectral-domain optical coherence tomography: The IN*OCT Consensus. Ophthalmology 2014;121:1572–1578. PMID 24755005

23. Sura AA, Chen L, Messinger JD, et al. Measuring the contributions of basal laminar deposit and Bruch’s membrane in age-related macular degeneration. Invest Ophthalmol Vis Sci 2020;61:19. PMID 33186466

24. Freund KB, Laud K, Lima LH, Spaide RF, Zweifel S, Yannuzzi LA. Acquired vitelliform lesions: correlation of clinical findings and multiple imaging analyses. Retina 2011;31:13–25. PMID 21102371

25. Tan ACS, Freund KB, Balaratnasingam C, Simhaee D, Yannuzzi LA. Imaging of pigment epithelial detachments with optical coherence tomography angiography. Retina 2018;38:1759–1769. PMID 29293207

26. Jakobiec FA, Barrantes PC, Yonekawa Y, Lad EM, Proia AD. Subretinal mononuclear cells in Coats’ disease studied with RPE65 and CD163: evidence for histiocytoid pigment epithelial cells. American Journal of Ophthalmology 2020;222:388–396. PMID 32950512

27. Curcio CA, Zanzottera EC, Ach T, Balaratnasingam C, Freund KB. Activated retinal pigment epithelium, an optical coherence tomography biomarker for progression in age-related macular degeneration. Invest Ophthalmol Vis Sci 2017;58:BIO211–BIO226. PMID 28785769

28. Sparrow JR, Marsiglia M, Allikmets R, et al. Flecks in recessive Stargardt disease: short-wavelength autofluorescence, near-infrared autofluorescence, and optical coherence tomography. Invest Ophthalmol Vis Sci 2015;56:5029–5039. PMID 26230768

29. Barros MH, Hauck F, Dreyer JH, Kempkes B, Niedobitek G. Macrophage polarisation: an immunohistochemical approach for identifying M1 and M2 macrophages. PLoS One 2013;8:e80908. PMID 24260507

30. Chistiakov DA, Killingsworth MC, Myasoedova VA, Orekhov AN, Bobryshev YV. CD68/macrosialin: not just a histochemical marker. Lab Invest 2017;97:4–13. PMID 27869795

31. Schaal KB, Freund KB, Litts KM, Zhang Y, Messinger JD, Curcio CA. Outer retinal tubulation in advanced age-related macular degeneration: optical coherence tomographic findings correspond to histology. Retina 2015;35:1339–1350. PMID 25635579

32. Etzerodt A, Moestrup SK. CD163 and inflammation: biological, diagnostic, and therapeutic aspects. Antioxid Redox Signal 2013;18:2352–2363. PMID 22900885

33. Karlstetter M, Scholz R, Rutar M, Wong WT, Provis JM, Langmann T. Retinal microglia: just bystander or target for therapy? Prog Retin Eye Res 2015;45:30–57. PMID 25476242

34. Lad EM, Cousins SW, Proia AD. Identity of pigmented subretinal cells in age-related macular degeneration. Graefes Arch Clin Exp Ophthalmol 2016;254:1239–1241. PMID 26728757

35. Margeta MA, Lad EM, Proia AD. CD163+ macrophages infiltrate axon bundles of postmortem optic nerves with glaucoma. Graefes Arch Clin Exp Ophthalmol 2018;256:2449–2456. PMID 30073622

36. Lamb TD, Pugh EN, Jr. Phototransduction, dark adaptation, and rhodopsin regeneration: the Proctor Lecture. Invest Ophthalmol Vis Sci 2006;47:5137–5152. PMID 17122096

37. Saari JC. Vitamin A and Vision. Subcell Biochem 2016;81:231–259. PMID 27830507

38. von Lintig J, Moon J, Babino D. Molecular components affecting ocular carotenoid and retinoid homeostasis. Prog Retin Eye Res 2021;80:100864. PMID 32339666

39. Xue Y, Shen SQ, Jui J, et al. CRALBP supports the mammalian retinal visual cycle and cone vision. J Clin Invest 2015;125:727–738. PMID 25607845

40. Liu T, Jenwitheesuk E, Teller DC, Samudrala R. Structural insights into the cellular retinaldehyde-binding protein (CRALBP). Proteins 2005;61:412–422. PMID 16121400

41. Ach T, Tolstik E, Messinger JD, Zarubina AV, Heintzmann R, Curcio CA. Lipofuscin re-distribution and loss accompanied by cytoskeletal stress in retinal pigment epithelium of eyes with age-related macular degeneration. Invest Ophthalmol Vis Sci 2015;56:3242–3252. PMID 25758814

42. Omri S, Omri B, Savoldelli M, et al. The outer limiting membrane (OLM) revisited: clinical implications. Clin Ophthalmol 2010;4:183–195. PMID 20463783

43. Polyak SL. The Retina. Chicago: University of Chicago; 1941.

44. Zanzottera EC, Ach T, Huisingh C, Messinger JD, Spaide RF, Curcio CA. Visualizing retinal pigment epithelium phenotypes in the transition to geographic atrophy in age-related macular degeneration. Retina 2016;36 Suppl 1:S12–S25. PMID 27658287

45. Matet A, Savastano MC, Rispoli M, et al. En face optical coherence tomography of foveal microstructure in full-thickness macular hole: a model to study perifoveal Muller cells. Am J Ophthalmol 2015;159:1142–1151 e1143. PMID 25728860

46. Lei J, Balasubramanian S, Abdelfattah NS, Nittala M, Sadda SR. Proposal of a simple optical coherence tomography-based scoring system for progression of age related macular degeneration. Graefes Arch Clin Exp Ophthalmol 2017;255:1551–1558. PMID 28534244

47. Nicolai U, Eckardt C. The occurrence of macrophages in the retina and periretinal tissues in ocular diseases. Ger J Ophthalmol 1993;2:195–201. PMID 8220098

48. Sheridan C, Hiscott P, Grierson I. Retinal pigment epithelium differentiation and dedifferentiation. In: Kirchhof B, Wong D (eds), Vitreo-retinal Surgery Essentials in Ophthalmology Berlin, Heidelberg: Springer; 2005.

49. Bonilha VL, Bell BA, Hu J, et al. Geographic atrophy: confocal scanning laser ophthalmoscopy, histology, and inflammation in the region of expanding lesions. Invest Ophthalmol Vis Sci 2020;61:15. PMID 32658960

50. Matsubara JA, Tian Y, Cui JZ, et al. Retinal distribution and extracellular activity of granzyme B: a serine protease that degrades retinal pigment epithelial tight junctions and extracellular matrix proteins. Front Immunol 2020;11:574. PMID 32318066

51. Chen KC, Jung JJ, Curcio CA, et al. Intraretinal hyperreflective foci in acquired vitelliform lesions of the macula: clinical and histologic study. Am J Ophthalmol 2016;164:89–98. PMID 26868959

52. Cherepanoff S, McMenamin PG, Gillies MC, Kettle E, Sarks SH. Bruch’s membrane and choroidal macrophages in early and advanced age-related macular degeneration. Br J Ophthalmol 2010;94:918–925. PMID 19965817

53. McLeod DS, Bhutto I, Edwards MM, Silver RE, Seddon JM, Lutty GA. Distribution and quantification of choroidal macrophages in human eyes with age-related macular degeneration. Invest Ophthalmol Vis Sci 2016;57:5843–5855. PMID 27802514

54. Elner SG, Elner VM, Nielsen JC, Torczynski E, Yu R, Franklin WA. CD68 antigen expression by human retinal pigment epithelial cells. Exp Eye Res 1992;55:21–28. PMID 1397126

55. Tatar O, Yoeruek E, Szurman P, et al. Effect of bevacizumab on inflammation and proliferation in human choroidal neovascularization. Arch Ophthalmol 2008;126:782–790. PMID 18541840

56. Sheridan CM, Pate S, Hiscott P, Wong D, Pattwell DM, Kent D. Expression of hypoxia-inducible factor-1alpha and −2alpha in human choroidal neovascular membranes. Graefes Arch Clin Exp Ophthalmol 2009;247:1361–1367. PMID 19590888

57. Ghazi-Nouri SM, Assi A, Limb GA, et al. Laser photocoagulation alters the pattern of staining for neurotrophin-4, GFAP, and CD68 in human retina. Br J Ophthalmol 2003;87:488–492. PMID 12642316

58. Simsek M, Ozates S, Gulpamuk B, Buyukeren B, Teke MY. Dedifferentiation of retinal pigment epithelium in a patient with chronic retinal detachment. Ophthalmic Surg Lasers Imaging Retina 2018;49:716–720. PMID 30222808

59. Josifovska N, Lumi X, Szatmari-Toth M, et al. Clinical and molecular markers in retinal detachment-From hyperreflective points to stem cells and inflammation. PLoS One 2019;14:e0217548. PMID 31185026

60. Sarks JP, Sarks SH, Killingsworth MC. Evolution of geographic atrophy of the retinal pigment epithelium. Eye 1988;2:552–577. PMID 2476333

61. Gocho K, Sarda V, Falah S, et al. Adaptive optics imaging of geographic atrophy. Invest Ophthalmol Vis Sci 2013;54:3673–3680. PMID 23620431

62. Penfold PL, Killingsworth MC, Sarks SH. Senile macular degeneration: the involvement of immunocompetent cells. Graefes Arch Clin Exp Ophthalmol 1985;223:69–76. PMID 2408968

63. Sennlaub F, Auvynet C, Calippe B, et al. CCR2(+) monocytes infiltrate atrophic lesions in age-related macular disease and mediate photoreceptor degeneration in experimental subretinal inflammation in Cx3cr1 deficient mice. EMBO Mol Med 2013;5:1775–1793. PMID 24142887

64. Schlereth SL, Kremers S, Schrodl F, Cursiefen C, Heindl LM. Characterization of antigen-presenting macrophages and dendritic cells in the healthy human sclera. Invest Ophthalmol Vis Sci 2016;57:4878–4885. PMID 27654414

65. Shirazi MF, Brunner E, Laslandes M, Pollreisz A, Hitzenberger CK, Pircher M. Visualizing human photoreceptor and retinal pigment epithelium cell mosaics in a single volume scan over an extended field of view with adaptive optics optical coherence tomography. Biomed Opt Express 2020;11:4520–4535. PMID 32923061

66. Lee B, Chen S, Moult EM, et al. High-speed, ultrahigh-resolution spectral-domain OCT with extended imaging range using reference arm length matching. Translational Vision Science & Technology 2020;9:12–12. PMID 32832219

67. Wu Z, Luu CD, Hodgson LAB, et al. Prospective longitudinal evaluation of nascent geographic atrophy in age-related macular degeneration. Ophthalmol Retina 2019. PMID 32088159

68. Dolz-Marco R, Litts KM, Tan ACS, Freund KB, Curcio CA. The evolution of outer retinal tubulation, a neurodegeneration and gliosis prominent in macular diseases. Ophthalmology 2017;124:1353–1367. PMID 28456420

69. Edwards MM, McLeod DS, Bhutto IA, Grebe R, Duffy M, Lutty GA. Subretinal glial membranes in eyes with geographic atrophy. Invest Ophthalmol Vis Sci 2017;58:1352–1367. PMID 28249091

70. Chen L, Messinger JD, Ferrara D, Freund KB, Curcio CA. Stages of drusen-associated atrophy in age-related macular degeneration visible via histologically validated fundus autofluorescence. Ophthalmology Retina 2020;11/30/20 online. PMID 33217617

71. Tan AC, Pilgrim M, Fearn S, et al. Calcified nodules in retinal drusen are associated with disease progression with age-related macular degeneration. Sci Transl Med 2018;10:466–477. PMID 30404862

72. Yang J, Antin P, Berx G, et al. Guidelines and definitions for research on epithelial-mesenchymal transition. Nat Rev Mol Cell Biol 2020;21:341–352. PMID 32300252

73. Gilmore AP. Anoikis. Cell Death Differ 2005;12 Suppl 2:1473–1477. PMID 16247493

74. Paoli P, Giannoni E, Chiarugi P. Anoikis molecular pathways and its role in cancer progression. Biochim Biophys Acta 2013;1833:3481–3498. PMID 23830918

75. Zhou M, Geathers JS, Grillo SL, et al. Role of epithelial-mesenchymal transition in retinal pigment epithelium dysfunction. Front Cell Dev Biol 2020;8:501. PMID 32671066

76. Guidry C, Medeiros NE, Curcio CA. Phenotypic variation of retinal pigment epithelium in age-related macular degeneration. Invest Ophthalmol Vis Sci 2002;43:267–273. PMID 11773041

77. Curcio CA, Millican CL. Basal linear deposit and large drusen are specific for early age-related maculopathy. Archives of Ophthalmology 1999;117:329–339. PMID 10088810

78. Ramrattan RS, van der Schaft TL, Mooy CM, de Bruijn WC, Mulder PGH, de Jong PTVM. Morphometric analysis of Bruch’s membrane, the choriocapillaris, and the choroid in aging. Invest Ophthalmol Vis Sci 1994;35:2857–2864. PMID 8188481

79. Mullins RF, Johnson MN, Faidley EA, Skeie JM, Huang J. Choriocapillaris vascular dropout related to density of drusen in human eyes with early age-related macular degeneration. Investigative Ophthalmology & Visual Science 2011;52:1606–1612. PMID 21398287

80. Seddon JM, McLeod DS, Bhutto IA, et al. Histopathological insights into choroidal vascular loss in clinically documented cases of age-related macular degeneration. JAMA Ophthalmol 2016;134:1272–1280. PMID 27657855

81. Lutty GA, McLeod DS, Bhutto IA, Edwards MM, Seddon JM. Choriocapillaris dropout in early age-related macular degeneration. Exp Eye Res 2020;107939. PMID 31987759

82. Balaratnasingam C, Yannuzzi LA, Curcio CA, et al. Associations between retinal pigment epithelium and drusen volume changes during the lifecycle of large drusenoid pigment epithelial detachments. Invest Ophthalmol Vis Sci 2016;57:5479–5489. PMID 27760262

83. Pan WW, Wubben TJ, Besirli CG. Photoreceptor metabolic reprogramming: current understanding and therapeutic implications. Commun Biol 2021;4:245. PMID 33627778

84. Hurley JB, Lindsay KJ, D. J. Glucose, lactate, and shuttling of metabolites in vertebrate retinas. J Neurosci Res 2015;93:1079–1092. PMID 25801286

85. Chinchore Y, Begaj T, Wu D, Drokhlyansky E, Cepko CL. Glycolytic reliance promotes anabolism in photoreceptors. Elife 2017;6. PMID 28598329

86. Lujan B, Roorda A, Knighton RW, Carroll J. Revealing Henle’s fiber layer using spectral domain optical coherence tomography. Invest Ophthalmol Vis Sci 2011;52:1486–1492. PMID 21071737

