## supplementary-material for "Hyperreflective foci, OCT progression indicators in age-related macular degeneration, include transdifferentiated retinal pigment epithelium"

### SUPPLEMENTARY MATERIALS

**Supplementary Methods: immunohistochemistry**

**Supplementary Figures**

Supplementary Figure 1. Henle fibers in the human macula.

Supplementary Figure 2. Positive and negative controls for immunohistochemistry

Supplementary Figure 3. Retinal pigment epithelium plumes exhibit directionality

Supplementary Figure 4. Duration and evolution of plumes

Supplementary Figure 5. Variations in RPE plume trajectory in atrophy.

Supplementary Figure 6. Gain- and loss-of-function of RPE and RPE-derived cells

Supplementary Figure 7. Dispersing melanosomes follow regionally specific trajectories of Müller glia

**Supplementary Tables**

Supplementary Table 1. Antibodies used

Supplementary Table 2: Morphologic phenotypes of RPE and its basal lamina in AMD

Supplementary Table 3. Demographics of clinic patients and study eyes

Supplementary Table 4. Plume disposition at final visit

Supplementary Table 5. Plume trajectory relative to HFL and macular quadrant

Supplementary Table 6. Plume association with atrophy

Supplementary Table 7. Characteristics of donor eyes

**Supplementary Methods: immunohistochemistry**

Cryosections were warmed, hydrated in phosphate-buffered saline (PBS) at room temperature, and processed for 15 minutes under steam with an antigen retrieval solution (Vector Labs, #H-3300-250) in glycerol. Quenched endogenous hydrogen peroxide with 3% $H_2O_2$ diluted from 30% $H_2O_2$ (Thermo Fisher Scientific Cat# H-325-100) with distilled $H_2O$ for ten minutes at room temperature. As recommended by the vendor, for CD68, this H2O2 quenching step was processed after primary antibody. Non-specific binding was blocked with 5% horse serum for 1 hour at room temperature, then incubated with primary antibodies of mouse anti-CD68, CD163, RPE65, or CRALBP at 4ºC for 40 hours. Following PBS wash, sections were incubated with avidin and biotin solutions (Vector Labs, #SP-2001) for 15 minutes respectively, washed (x3), then incubated with horse anti-mouse secondary antibody (Vector Labs Vectastain Elite ABC Kit RTU Universal, #PK-7200), washed (x3), and incubated with ABC complex solution (Vector Labs Vectastain Elite ABC Kit RTU Universal, #PK-7200) for 30 minutes at room temperature, and washed. A red reaction product was developed with Vector AEC Peroxidase Substrate Kit (Vector Labs, Burlingame CA USA #SK-4200) to the desired color (~30 minutes). After washing with distilled water (5 minutes x 3), sections were counterstained with hematoxylin solution for 5 minutes followed by 20 seconds bluing solution (Poly Scientific R&D Corp, Bay Shore NY USA; #k047) then washed. Coverslips were mounted with aqueous medium (VectaMount AQ, Vector Labs, #H-5501-60) and sealed with nail polish, air-dried, and stored at 4ºC.

**Supplementary figures**

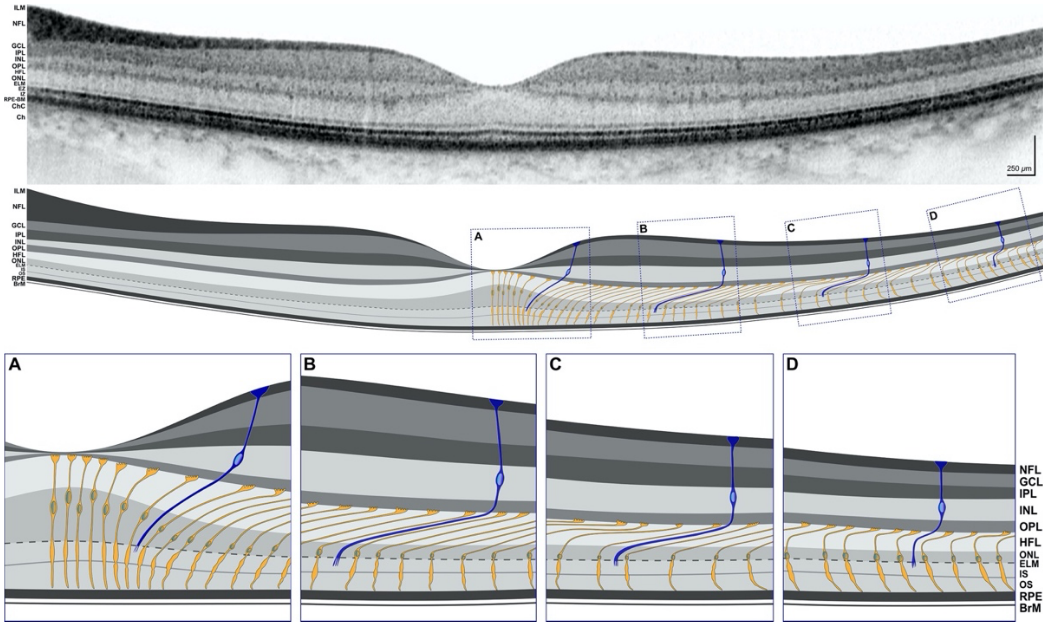


***<u>Supplementary Figure 1. Henle fibers in the human macula.</u>***

***Top**:* Foveal OCT B-scan of a healthy female aged between 66 and 70 years demonstrating reflective bands corresponding to retinal layers. In reference to the middle panel, the Henle fiber layer is visible, especially under the fovea. ***Middle:*** Schematized OCT scan shows retinal layers with cone photoreceptors *(*yellow*)* and Müller glia (blue). Müller glia span the retina, terminating at the external limiting membrane; for clarity, their branches in plexiform layers are omitted. ***Bottom**:* (**A**) At the foveal center, Henle fibers are short and perpendicular*. (***B**) In the parafovea, Henle fibers are long and oblique. (**C**) In the perifovea, Henle fibers gradually shorten. (**D**) At the macula edge, Henle fibers are very short and nearly vertical. Henle fibers are not present elsewhere in the retina. Schematic of B-scan was prepared (Illustrator, version 23.0.3, Adobe Inc., San Jose, CA) from an OCT B-scan (Spectralis, Heidelberg Engineering, Germany). Schematic of cellular trajectories was prepared in reference to a histological section of a donor eye (female, 76-80 years) immuno-labeled for glial fibrillary acidic protein in Müller glia.[87]

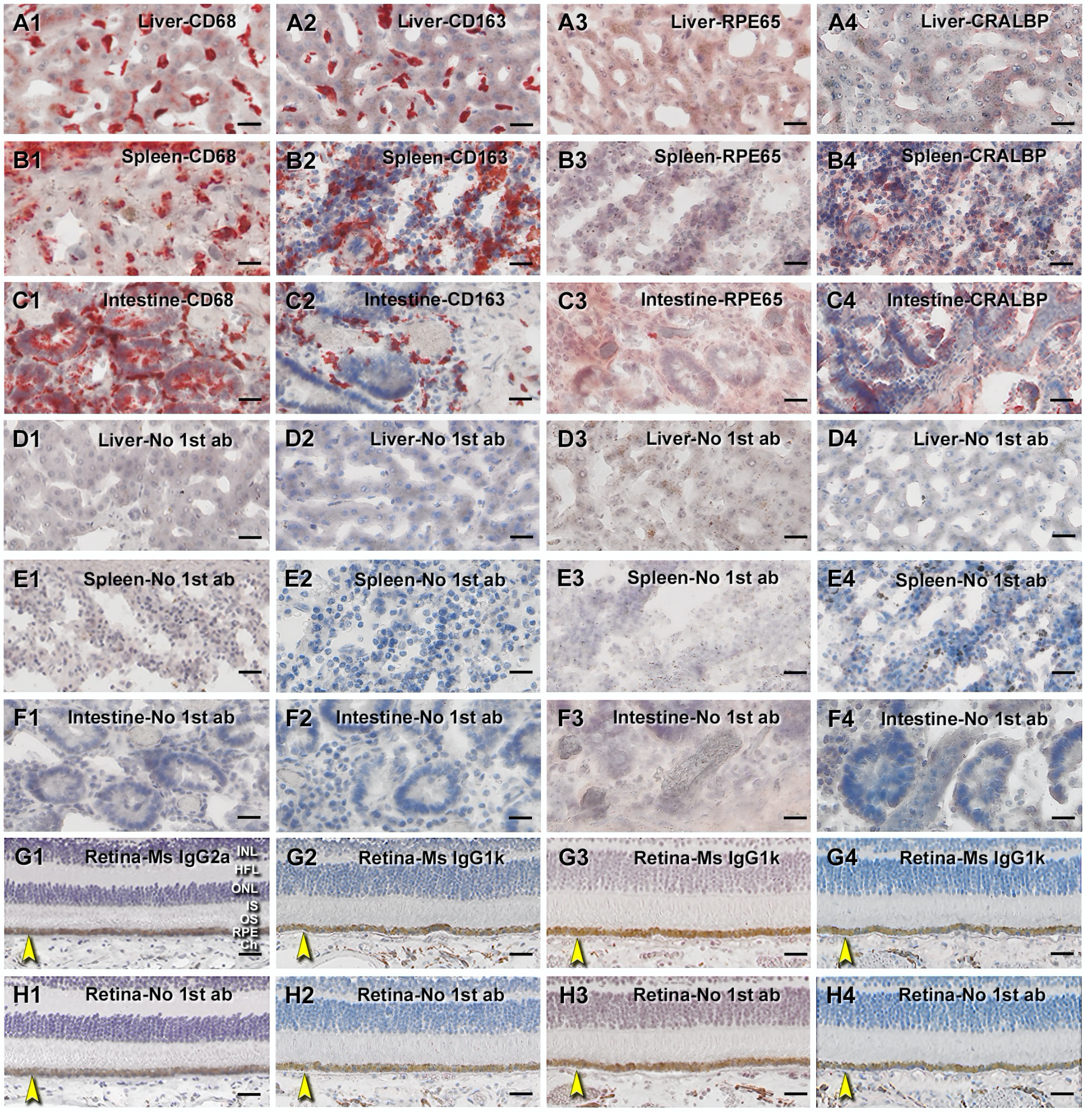


***Supplementary Figure 2. Positive and negative controls for immunohistochemistry.***

A cryosection of human tissue array with liver, spleen, and intestine tissue made in-house was used as positive and negative controls for immunohistochemistry studies with antibodies to CD68, CD163, RPE65, and CRALBP (positive A to C, negative D to H). Human retinal sections from an unremarkable eye (#3, Supplementary Table 7) was used for isotype and sham controls. BrM was pointed by yellow arrows. Scale bars are

20 µm for non-retinal tissues and 50 µm for retina. (**A1** to **A4**) CD68+, CD163+, RPE65-, and CRALBP- Kupffer cells in liver tissue. (**B1** to **B4**) CD68+, CD163+, RPE65-, and CRALBP+ red pulp cells in spleen tissue. (**C1** to **C4**) CD68+, CD163+, RPE65+, and CRALBP+ cells in intestine tissue. (**D1** to **D4**) Liver sham controls. (**E1** to **E4**) Spleen sham controls. (**F1** to **F4**) Intestine sham controls. (**G1** to **G4**) Retina isotype controls with mouse IgG2a for CD68 and mouse IgG1k for CD163, RPE65, and CRALBP. (**H1** to **H4**) Retina sham controls for CD65, CD163, RPE65, and CRALBP. All human tissue sections have CD68 and CD163 immunoreactivities and uncertain RPE65 and CRALBP immunoreactivities. All isotype and sham controls omitting primary antibodies control are negative.

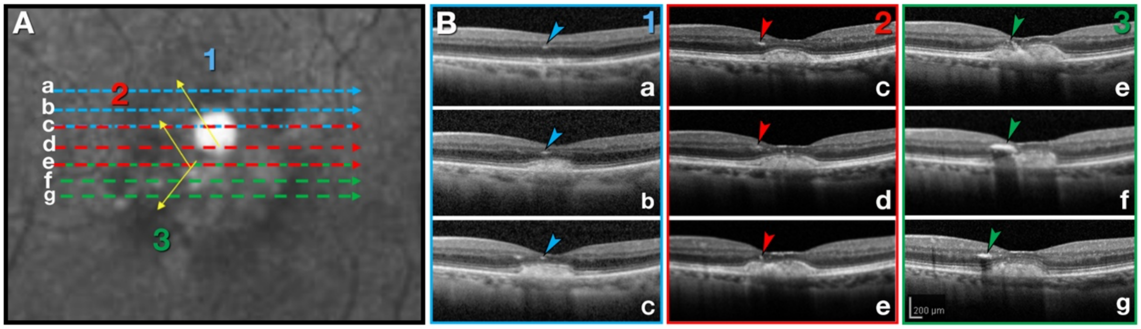


***<u>Supplementary Figure 3. Retinal pigment epithelium plumes exhibit directionality.</u>***

RPE plumes radiate outward from the fovea and thus may track as small reflective features across sequential horizontally oriented optical coherence tomography B-scans. **(A)** Near-infrared reflectance (NIR) image shows plumes 1-3, with arrows indicating plume direction into the retina. Dashed lines (blue, red, green) correspond to scan lines a-g in panel B1-3. Positions of plumes, which are not visible in NIR, were assigned to this image and corresponded to the B-scans with the device software. (**B)** Blue, red and green arrowheads indicate plumes 1-3, transected by each B-scan. In B1-c, a second plume is visible in addition to tracked plume 1. Scale bar in g applies to all panels of B.

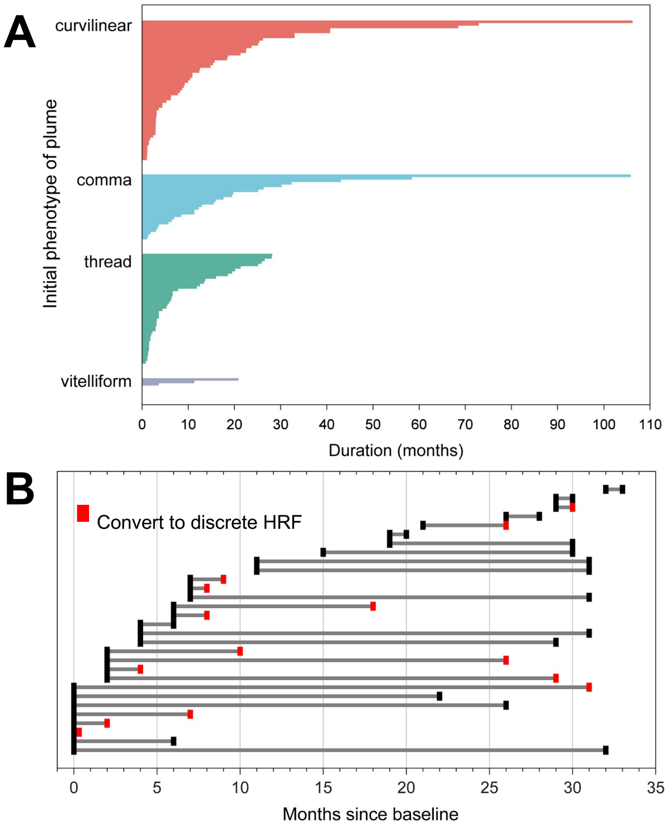


***<u>Supplementary Figure 4. Duration and evolution of plumes.</u>***

(**A**) Duration of each of 129 plumes, stratified by morphology, and displayed in descending order of duration. The longest lasting plumes were curvilinear and comma. (**B**) Each plume in one eye with the most plumes represented as a single horizontal line. Duration is shown, with conversion to discrete HRF, no longer associated with the RPE, indicated; representative of data from 3 eyes.

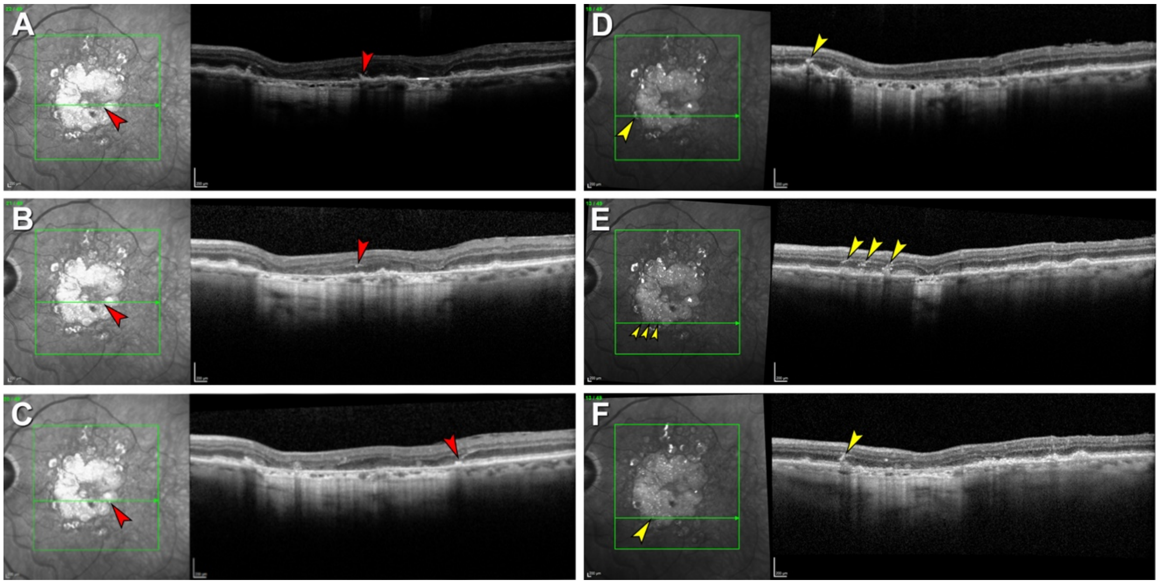


***Supplementary Figure 5. Variations in RPE plume trajectory in atrophy.***

Plumes located along the border of complete RPE and outer retinal atrophy (cRORA, [1]) are indicated in the near infrared reflectance images at the left of each panel. Green lines indicate areas covered by OCT volumes and locations of individual B-scans in A-F. (**A** and **B**) Posterior (A) and anterior (B) components of a plume (red arrowheads) traversing radially away from the fovea, i.e., congruent with the HFL. (**C**) Another plume (red arrowhead) is congruent with the normal HFL, adjacent to a hyporeflective wedge [2]. (**D** to **F**) Examples of plumes traversing anteriorly in the opposite direction, i.e., non-congruent with the normal HFL (yellow arrowheads), near the border of atrophy, possibly due to subsidence (sinking) of outer retinal layers in atrophy.

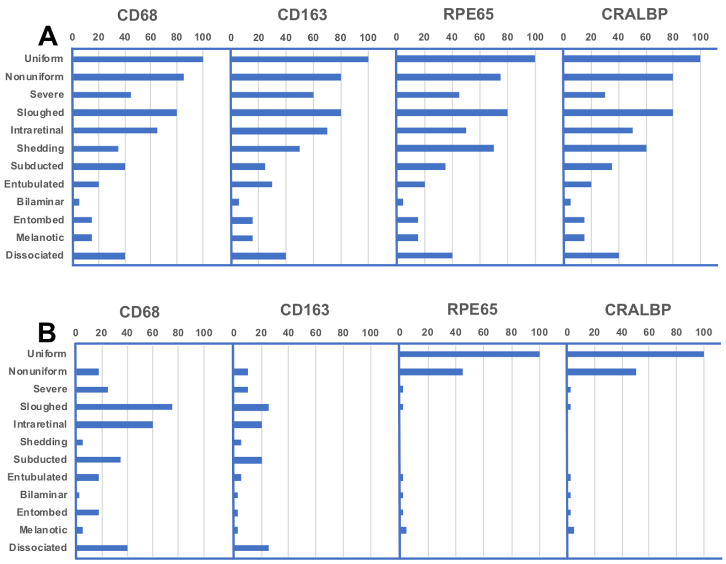


***Supplementary Figure 6. Gain- and loss-of-function of RPE and RPE-derived cells.***

(**A**) The distribution of phenotypes (vertical axis) was similar across the sections used for immunohistochemistry for retinoid markers RPE65 and CRALBP and immune markers CD68 and CD163 (at top of graphs). Uniform and Non-uniform phenotypes are age normal. Of the others, Entubulated and Subducted are found in geographic atrophy and neovascular AMD, Melanotic is found only in neovascular AMD, and others are found at all AMD diagnostic stages. Vitelliform and Vacuolated are not shown due to small numbers in the current sample. (**B**) Bar lengths represent the percentage labelled (scoring with 0-not labelled, 1-some labelled, and 2-all abnormal cells labelled; total scores/ maximum possible score = 40 x 100) by CD68, CD163, RPE65 and CRALBP. Age-normal uniform cells are strongly labelled by retinoid markers and are not labelled by immune markers. Non-uniform cells exhibit a reduced proportion of retinoid markers and a small proportion of immune markers. Phenotypes below “severely non-uniform” on the vertical axis are considered abnormal. These are labeled for immune markers (up to 75%) and minimally labelled for retinoid markers (<5%), especially “Sloughed” and “Intraretinal” morphologies corresponding to HRF in OCT.

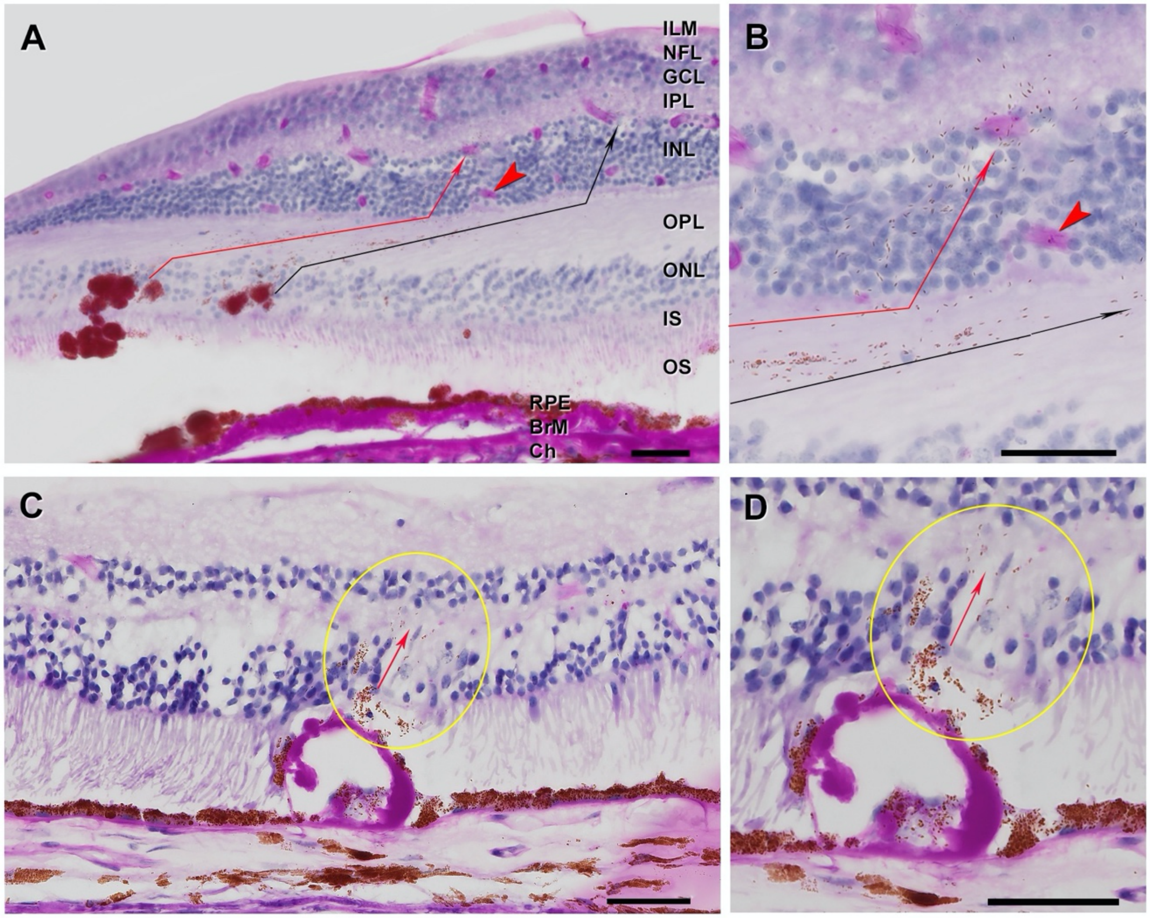


***Supplementary Figure 7. Dispersing melanosomes follow regionally specific trajectories of Müller glia.***

A and B, donor aged 96-100 years (#17, Supplementary Table 7). C and D, donor aged 81-85 years (#1, Supplementary Table 7). All scale bars are 50 µm. Cryosection, periodic acid Schiff hematoxylin stain. (**A** and **B**) In the macula RPE detritus follows the obliquely oriented HFL fibers and crosses vertically into the INL. Thin red and black arrows indicate detritus emanating from two groups of cells in the ONL. (**C** and **D)** In the periphery where HFL is absent, RPE organelles enter vertically retina directly above an artifactually empty druse lined by BLamD and lacking overlying RPE.

**Supplementary tables**

***Supplementary Table 1: Antibodies used***

| Antibody Name | Clone # | Cat# | Lot# | Isotype | Dilution | Vendor Web Link |
|---|---|---|---|---|---|---|
| CD68 Monoclonal | 514H12 | MA1-80133 | C2619 | Mouse IgG 2a | 1:100 | https://www.fishersci.com/shop/products/anti-cd68-clone-514h12/ma180133 |
| CD163 Monoclonal | 10D6 | MA5-11458 | D1118 | Mouse IgG 1 | 1:100 | https://www.fishersci.com/shop/products/anti-cd163-clone-10d6-thermo-scientific-pierce/pima511458 |
| RPE65 Monoclonal | 401.8B11.3D9 | MA1-16578 | UE2776581 | Mouse IgG 1k | 1:150 | https://www.fishersci.com/shop/products/anti-rpe65-clone-401-8b11-3d9/ampa6578 |
| CRALBP Monoclonal | B2 | MA1-813 | UB272117 | Mouse IgG 1 | 1:500 | https://www.fishersci.com/shop/products/anti-cralbp-clone-b2/ma1813# |
| Mouse IgG 2a | PPV-04 | MA1-10418 | UC282768 | Mouse IgG 2a | 1:1000 | https://www.fishersci.com/shop/products/mouse-igg2a-clone-ppv-04-isotype-control-thermo-scientific-pierce/pima110418 |
| Mouse IgG 1Kappa | P3.6.2.8.1 | 14-4714-82 | 1957953 | Mouse IgG 1k | RTU | https://www.fishersci.com/shop/products/igg1-k-mouse-clone-p3-6-2-8-1-isotype-control-ebioscience-3/501129668 |
| Horse anti-Mouse Biotin | NA | PK-7200 | ZE0720 | Horse IgG | RTU | https://vectorlabs.com/catalogsearch/result/?q=pk-7200 |
| RTU, ready to use | | | | | | |

***Supplementary Table 2: Morphologic phenotypes of RPE and its basal lamina in AMD***

| Name | Attached to basal lamina | Description (in toluidine-blue stained sub-micrometer sections of osmium-tannic-acid paraphenylenediamine post-fixed human eyes) |
|---:|:---:|---|
| Uniform | Yes | Uniform height and pigmentation; hexagonal |
| Non uniform | Yes | Slightly non-uniform morphology and pigmentation; hexagonal |
| Very non uniform | Yes | Very non-uniform morphology and pigmentation |
| Sloughed | No | Continuous epithelium with equally pigmented spherical cells sloughed into sub-retinal space |
| Shedding | Yes | Continuous epithelium with groups of organelles (non-nucleated, non-membrane bound) shed into thick underlying basal laminar deposits; granule aggregates form in the cell bodies. |
| Bilaminar | Yes | Double layers; usually neovascular AMD |
| Vacuolated | Yes | A single large vacuole, sometimes with contents, delimited apically by extremely effaced cytoplasm (rare) |
| Intraretinal | No | Anterior migration of nucleated and pigmented cells, singly and in groups, through the external limiting membrane; mostly spherical, some amoeboid |
| Vitelliform | No | Massively liberated RPE organelles in the subretinal space |
| Dissociated | No | Spherical cells in scattered in atrophic areas, usually adherent to persistent BLamD. |
| Entombed | Yes | Entombed by fibrovascular scar in neovascular AMD, intermingled with other cells and fluid in the same layer |
| Subducted | No | Rounded or flattened, in sub-RPE-basal laminar space, with varying numbers of RPE-characteristic organelles |
| Melanotic | Yes | Single or multiple cells in layers with black spherical melanosomes, some large (up to 5 µm diameter), in sub-retinal or sub-RPE space; neovascular AMD only |
| Entubulated | No | In lumen of outer retinal tubulation (tubes of photoreceptors, mainly cones, scrolled by Müller glia) |
| Atrophy with BLamD | No | No RPE, persistent basal laminar deposits |
| Atrophy without BLamD | No | No RPE, no basal laminar deposit (retina consists of gliotic Müller cells in contact with inner collagenous layer of Bruch's membrane |

Phenotypes defined using 1-µm-thick epoxy resin sections of osmium-tannic-acid paraphenylenediamine post-fixed human eyes with AMD, stained with toluidine blue, referenced to previously published AMD morphology [3-5]. All cells except where noted are nucleated and have abundant lipofuscin, melanolipofuscin, and characteristic spindle-shaped melanosomes in the cell bodies. Melanosomes are abundant in apical processes [6], which may be lost due to disease or to artifactual detachment of photoreceptors from RPE or both.

Lipofuscin and melanolipofuscin are ovoid (~1 µm in diameter), densely packed, and green-bronze-olive color in toluidine blue stain, depending on the specimen. Cell morphologies recognized in this preparation are readily identified in the cryosections used in this study. Granule aggregates (Shedding phenotype) are also visible in RPE cell bodies by fluorescence microscopy [7]. Vitelliform [8] is under active investigation [9] and was not identified in the current series but is included for completeness.

***Supplementary Table 3. Demographics of clinic patients and study eyes***

| **Variable (SD, standard deviation)** | **Value** |
|---|---|
| Number of patients | 44 |
| Number of eyes | 61 |
| Age, mean ± standard deviation (years) | 79.4 ± 7.7 |
| Gender: Female, N (%) | 29 (47.5) |
| Eyes with RPE plume | 29 (47.5) |
| Follow-up duration (years, per individual eyes) | 4.7 ± 0.09 |
| Total number of plumes | 129 |
| Plumes/eye (range, SD) | 4.0 (1-30, 5.9) |
| Distance in µm, fovea to plume center, mean (SD); range | 1555 (SD 876); 0 – 3558 |
| **Presence of AMD features: N (%)** | |
| Soft drusen only | 10 (16.4) |
| Cuticular drusen only | 8 (13.1) |
| Mixed drusen types | 34 (55.7) |
| Drusenoid pigment epithelial detachment* | 27 (44.3) |
| Complete RPE and outer retinal atrophy** | 29 (47.5) |
| *≥350 µm greatest linear diameter [10]<br>**cRORA as defined [1] | |

***Supplementary Table 4. Plume disposition at final visit***

| **Fates of 129 individual plumes (baseline vs final)** | **# (% of 129)** |
|---|---|
| Complete resolution *with conversion* to another plume morphology or discrete HRF | 38 (29.5) |
| Complete resolution *without conversion* to another plume morphology or discrete HRF | 36 (27.9) |
| Unresolved *with conversion* to another plume morphology or discrete HRF | 35 (27.1) |
| Unresolved *without conversion* to another plume morphology or discrete HRF | 20 (15.5) |
| **Total** | **129 (100)** |

***Supplementary Table 5. Plume trajectory relative to HFL and macular quadrant***

| | **Congruent With normal HFL** | | **Non-congruent With normal HFL** | |
|---|---|---|---|---|
| | **# eyes** | **# plumes** | **# eyes** | **# plumes** |
| Curvilinear | 17 | 42 | 7 | 15 |
| Comma | 14 | 19 | 2 | 6 |
| Thread | 12 | 36 | 5 | 7 |
| Vitelliform | 3 | 4 | 0 | 0 |
| Superior temporal | 13 | 34 | 0 | 0 |
| Superior nasal | 15 | 23 | 4 | 19 |
| Inferior nasal | 9 | 18 | 2 | 5 |
| Inferior temporal | 17 | 26 | 3 | 4 |

***Supplementary Table 6. Plume association with atrophy***

| **Association with atrophy (at final visit)** | **# (% of 129 total)** |
|---|---|
| Associated with cRORA | 73 (56.6) |
| Congruent | 48 (37.2) |
| Incongruent | 25 (19.4) |
| At cRORA border | 16 (12.4) |
| Associated with iRORA | 6 (4.7) |
| Congruent | 6 (4.7) |
| Incongruent | 0 (0) |
| Not associated with cRORA or iRORA | 50 (38.8) |
| **Total** | **129** |

cRORA, complete RPE and outer retinal atrophy; iRORA, incomplete RPE and outer retinal atrophy [1, 11]
Congruent, incongruent; relative to trajectory of normal HFL at that location

***Supplementary Table 7. Characteristics of donor eyes***

| Order | Age range | Sex | D-P, hours | Histologic diagnosis |
|---|---|---|---|---|
| 1 | 81-85 | M | 4.05 | Unremarkable |
| 2 | 86-90 | F | 5.48 | Unremarkable |
| 3 | 76-80 | F | 4.82 | Unremarkable |
| 4 | 91-95 | F | 3.75 | Unremarkable |
| 5 | 86-90 | F | 4.75 | Early-intermediate AMD |
| 6 | 86-90 | F | 3.25 | Early-intermediate AMD |
| 7 | 91-95 | M | 3.63 | Early-intermediate AMD |
| 8 | 86-90 | F | 4.95 | Early-intermediate AMD |
| 9 | 81-85 | F | 3.58 | Early-intermediate AMD |
| 10 | 96-100 | F | 3.98 | Early-intermediate AMD |
| 11 | 96-100 | F | 3.12 | Early-intermediate AMD |
| 12 | 91-95 | M | 3.45 | Atrophic AMD |
| 13 | 91-95 | F | 3.78 | Atrophic AMD |
| 14 | 81-85 | F | 3.50 | Atrophic AMD |
| 15 | 86-90 | F | 3.42 | Atrophic AMD |
| 16 | 86-90 | M | 4.15 | Atrophic AMD |
| 17 | 96-100 | F | 2.33 | Atrophic AMD |
| 18 | 86-90 | F | 3.50 | Neovascular AMD |
| 19 | 91-95 | F | 4.15 | Neovascular AMD |
| 20 | 86-90 | F | 3.80 | Neovascular AMD |

Notes:
Total 20 eyes of 20 white donors. Age ranges in years: 76-80; 81-85; 86-90; 91-95; 96-100; mean age 89.2 ± 5.0 years; 4 males and 16 females. D-P, death to processing time in hours; mean D-P 3.87 ± 0.72 hours; 4 unremarkable, 7 early-intermediate AMD, 6 atrophy AMD, 3 neovascular AMD.

### References

1. Sadda SR, Guymer R, Holz FG, et al. Consensus definition for atrophy associated with age-related macular degeneration on optical coherence tomography: CAM Report 3. *Ophthalmology* 2018;125:537-548. PMID 29103793
2. Monés J, Biarnes M, Trindade F. Hyporeflective wedge-shaped band in geographic atrophy secondary to age-related macular degeneration: an underreported finding. *Ophthalmology* 2012;119:1412-1419. PMID 22440276
3. Zanzottera EC, Messinger JD, Ach T, Smith RT, Freund KB, Curcio CA. The Project MACULA retinal pigment epithelium grading system for histology and optical coherence tomography in age-related macular degeneration. *Invest Ophthalmol Vis Sci* 2015;56:3253-3268. PMID 25813989
4. Zanzottera EC, Messinger JD, Ach T, Smith RT, Curcio CA. Subducted and Melanotic cells in advanced age-related macular degeneration are derived from retinal pigment epithelium. *Invest Ophthalmol Vis Sci* 2015;56:3269-3278. PMID 26024109
5. Curcio CA, Zanzottera EC, Ach T, Balaratnasingam C, Freund KB. Activated retinal pigment epithelium, an optical coherence tomography biomarker for progression in age-related macular degeneration. *Invest Ophthalmol Vis Sci* 2017;58:BIO211-BIO226. PMID 28785769
6. Pollreisz A, Neschi M, Sloan KR, et al. An atlas of human retinal pigment epithelium organelles significant for clinical imaging. *Invest Ophthalmol Vis Sci* 2020;61:13. PMID 32648890
7. Ach T, Tolstik E, Messinger JD, Zarubina AV, Heintzmann R, Curcio CA. Lipofuscin re-distribution and loss accompanied by cytoskeletal stress in retinal pigment epithelium of eyes with age-related macular degeneration. *Invest Ophthalmol Vis Sci* 2015;56:3242-3252. PMID 25758814
8. Arnold JJ, Sarks JP, Killingsworth MC, Kettle EK, Sarks SH. Adult vitelliform macular degeneration: a clinicopathological study. *Eye* 2003;17:717-726. PMID 12928683
9. Chen KC, Jung JJ, Curcio CA, et al. Intraretinal hyperreflective foci in acquired vitelliform lesions of the macula: clinical and histologic study. *Am J Ophthalmol* 2016;164:89-98. PMID 26868959
10. Cukras C, Agron E, Klein ML, et al. Natural history of drusenoid pigment epithelial detachment in age-related macular degeneration: Age-Related Eye Disease Study Report No. 28. *Ophthalmology* 2010;117:489-499. PMID 20079925
11. Guymer RH, Rosenfeld PJ, Curcio CA, et al. Incomplete retinal pigment epithelial and outer retinal atrophy (iRORA) in age-related macular degeneration: CAM Report 4. *Ophthalmology* 2020;127:394-409. PMID 31708275
